# Association between anaemia, micronutrient status, and pneumococcal vaccine responses in young Kenyan children

**DOI:** 10.64898/2026.07.03.26357225

**Authors:** Kelvin Mokaya Abuga, Henry K Karanja, Katherine Gallagher, Bridgious Walusimbi, Beth Wanjiku Mugure, Caren Konah Koli, Beryl Masinde, Timothy Etyang, Angela Karani, Elizabeth M Indeje, John Muthii Muriuki, Laura Hammitt, Samson Kinyanjui, Calman A MacLennan, Manfred Nairz, J Anthony G Scott, Alison M Elliott, Gyaviira Nkurunungi, Sarah H Atkinson

## Abstract

Anaemia and micronutrient deficiencies are common in low- and middle-income countries, but whether nutritional status influences pneumococcal vaccine immunogenicity in children remains poorly characterised. We examined associations between pneumococcal vaccine responses in 670 Kenyan children enrolled in three vaccine trials: PRISM (PCV10; n=195; NCT01028326), FPCV (fractional- and full-dose PCV10/PCV13; n=306; NCT03489018), and PATH-wSP (whole-cell pneumococcal vaccine; n=169; NCT02543892) and baseline anaemia and micronutrient status (iron, folate, zinc, and vitamins A, B12, D, and E). Analyses were performed separately for each trial. In the 1 mg PATH-wSP group, anaemia and iron deficiency anaemia were associated with lower composite IgG responses (adj. β −0.8 [−1.2, −0.4] and adj. β −0.7 [−1.2, −0.2], respectively). Higher vitamin B12 concentrations were associated with higher composite IgG (adj. β 0.4 [0.1, 0.7]) and opsonophagocytic activity (adj. β 0.4 [0.2, 0.7]) responses in the PRISM trial. There was little evidence of associations between biomarkers of iron status, folate, zinc, or vitamins A, D, and E and pneumococcal vaccine responses. These findings suggest that nutritional status is not a uniform determinant of pneumococcal vaccine immunogenicity, but that specific nutritional factors, particularly anaemia and vitamin B12 status, may influence vaccine responses in a context-dependent manner.

## Introduction

*Streptococcus pneumoniae* (pneumococcus) is a leading cause of hospitalisation and death, accounting for an estimated 829,000 deaths globally in 2019.^1^ A substantial proportion of pneumococcal deaths occur in young children.^2^ Although pneumococcal conjugate vaccines (PCVs) have markedly reduced vaccine-serotype carriage and invasive pneumococcal disease,^3,4^ morbidity and mortality remain high in low-resource settings.^2^ Vaccine-induced immune responses are often suboptimal in these settings.^5^ At the level of vaccine coverage in 2013, it was estimated that 10 million children were vaccinated but remained unprotected against invasive pneumococcal disease,^6^ underscoring the urgent need to identify modifiable determinants of suboptimal vaccine responses.

Anaemia and micronutrient deficiencies are highly prevalent in low-resource settings,^7,8^ where they may contribute to suboptimal vaccine-induced immune responses and increase the burden of pneumococcal disease.^9–11^ Several micronutrients regulate key innate and adaptive immune pathways involved in vaccine responses.^12,13^ However, their relationship with pneumococcal vaccine immunogenicity remains poorly characterised, with evidence limited to a small number of heterogeneous studies.^14–16^ Associations may vary by micronutrient, vaccine platform, and serotype. For example, anaemia and iron deficiency were associated with reduced PCV10 (Synflorix^®^) responses to serotype 19,^14^ while higher vitamin B12 concentrations were associated with stronger responses to the 23-valent pneumococcal polysaccharide vaccine (Pneumovax^®)^)^15^. In contrast, no associations were observed between haemoglobin or micronutrient status (zinc, vitamin A and vitamin C) and responses to Pneumovax^®^.^16^ Whether these differences reflect biological heterogeneity or variation in age, study design, and vaccine platform remains uncertain.

We investigated the associations of baseline anaemia and micronutrient status (iron, folate, zinc, and vitamins A, B12, D, and E) with post-vaccination antibody responses across three pneumococcal vaccine trials in Kenyan children, spanning different ages, vaccine platforms, and dosing regimens.

## Results

### Characteristics of study participants

We included 670 children across three pneumococcal vaccine trials (Figure 1). The cohorts differed substantially in study design (Table 1) and participant characteristics (Table 2).

**Figure 1.**
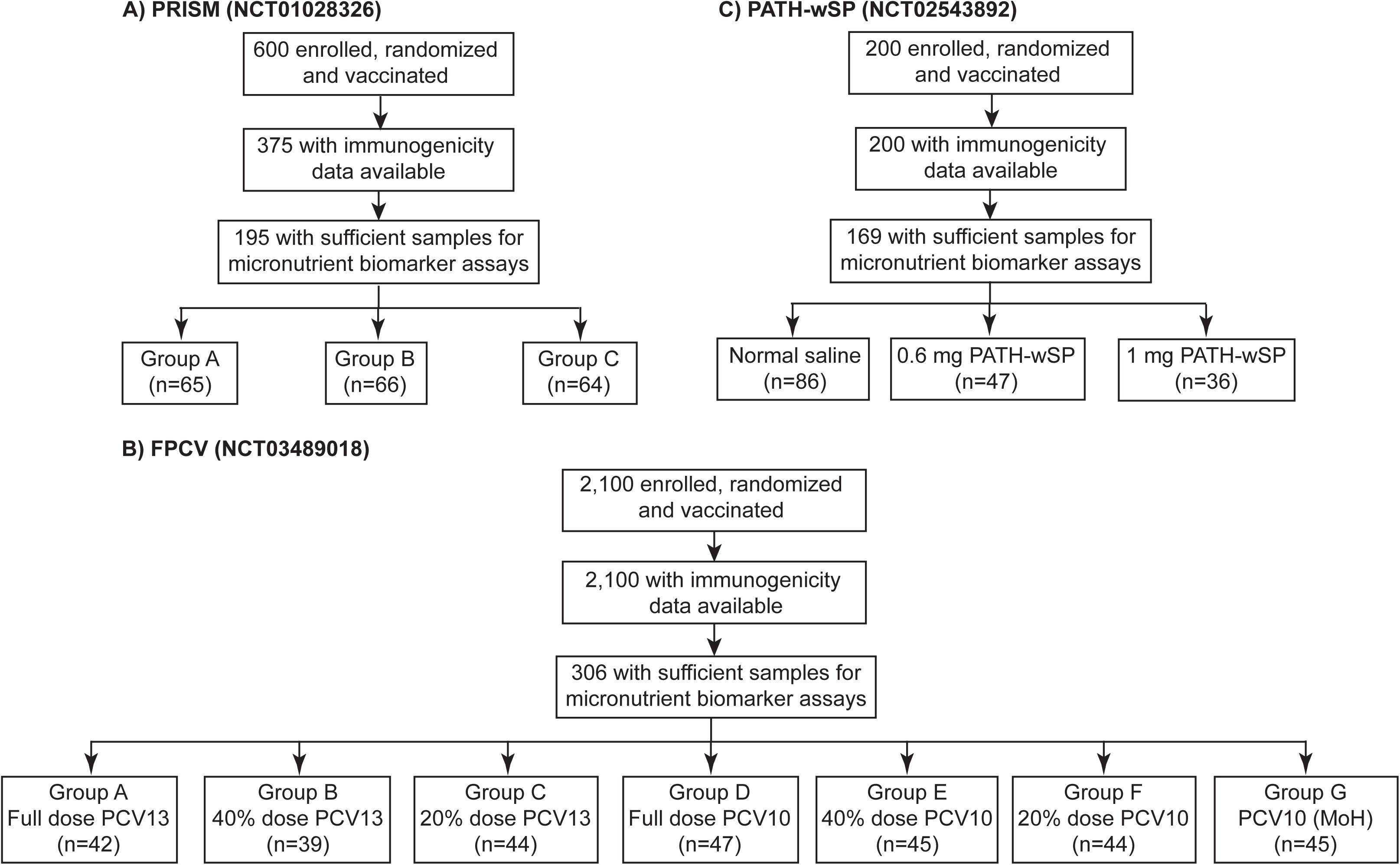
Selection of participants enrolled across three clinical trials. A) PRISM (NCT01028326): Participants received two doses of PCV10 vaccine administered either 60 days apart (Group A) or 180 days apart (Group B), or control vaccines (Group C). B) FPCV (NCT03489018): Participants received PCV13 at full dose (Group A), 40% dose (Group B), or 20% dose (Group C), or PCV10 at full dose (Group D), 40% dose (Group E), or 20% dose (Group F); two primary doses at 6 weeks and 14 weeks of age, and a booster at 9 months of age. A comparator group (Group G) received full-dose PCV10 according to the Kenyan Ministry of Health (MoH) immunisation schedule (3P+0) at 6, 10, and 14 weeks of age. C) PATH-wSP (NCT02543892): Participants received two doses of 0.6 mg or 1 mg of the whole-cell pneumococcal vaccine or placebo (normal saline), with vaccines administered 28 days apart.

**Table 1.** Characteristics of three pneumococcal vaccine trials in Kenyan children.

|  | **PRISM (n=195)** | **FPCV (n=306)** | **PATH-wSP (n=169)** |
| --- | --- | --- | --- |
| Study period | January – September 2010 | March 2019 – November 2021 | August 2016 – January 2018 |
| Age at vaccination | 12–59 months | 6–8 weeks | 12–21 months |
| Vaccine | PCV10 | PCV10 (20%, 40%, and full dose); PCV13 (20%, 40%, and full dose) | PATH-wSP |
| Vaccine schedule | 2 doses<br>**Group A:** PCV10 (day 0), PCV10 (day 60), DTaP (day 180)<br>**Group B:** PCV10 (day 0), DTaP (day 60), and PCV10 (day 180) | 2 primary doses (6 and 14 weeks of age) + booster (9 months of age)<br>**Group A:** full dose PCV13<br>**Group B:** 40% dose PCV13<br>**Group C:** 20% dose PCV13<br>**Group D:** full dose PCV10<br>**Group E:** 40% dose PCV10<br>**Group F:** 20% dose PCV10 | 2 doses (days 0 and 56) of **0.6 mg** PATH-wSP or **1 mg** of the PATH-wSP |
| Control group | **Group C:** HAV (day 0), DTaP (day 60), and HAV (day 180) | **Group G:** Full-dose PCV10 as per the Kenyan Ministry of Health immunisation schedule (3P+0 schedule) | **Placebo** (normal saline) |
| Blood sampling timepoints | Baseline; 30 days after first vaccination; and 30 days after second vaccination (Group A and C) or 30 days after third vaccination (Group B) | Baseline; 4 weeks after primary doses; 4 weeks after booster | Baseline; 4 weeks after second dose |
| Nutritional biomarkers | Ferritin, serum iron, transferrin, folate, zinc, and vitamins A, B12, D, and E | Haemoglobin, ferritin, serum iron, transferrin, folate, zinc, and vitamins A, B12, and E | Haemoglobin, ferritin, serum iron, transferrin, folate, zinc, and vitamins A, B12, and E |
| Vaccine response outcomes (serotypes/antigens) | 1, 4, 5, 6B, 7F, 9V, 14, 18C, 19F, and 23F | **PCV10:** 1, 4, 5, 6B, 7F, 9V, 14, 18C, 19F, and 23F<br>**PCV13:** 1, 3, 4, 5, 6A, 6B, 7F, 9V, 14, 18C, 19A, 19F, and 23F | L460D, PspA-Fam1, PhtD, BCH0785, StkP, PcpA, SPWCV, PiuA, PiaA |
| Outcome measures | IgG, OPA | IgG | IgG |
| Outcome timepoints | 30 days after first vaccination | 28 days after primary doses | 28 days after two doses |

**Abbreviations:** PCV10 denotes 10-valent pneumococcal conjugate vaccine; PCV13, 13-valent pneumococcal conjugate vaccine; HAV, hepatitis A vaccine; DTaP, diphtheria–tetanus–acellular pertussis vaccine; IgG, immunoglobulin G; OPA, opsonophagocytic activity; 2P+1, two primary doses plus booster; 3P+0, three primary doses without booster.

**Table 2.**
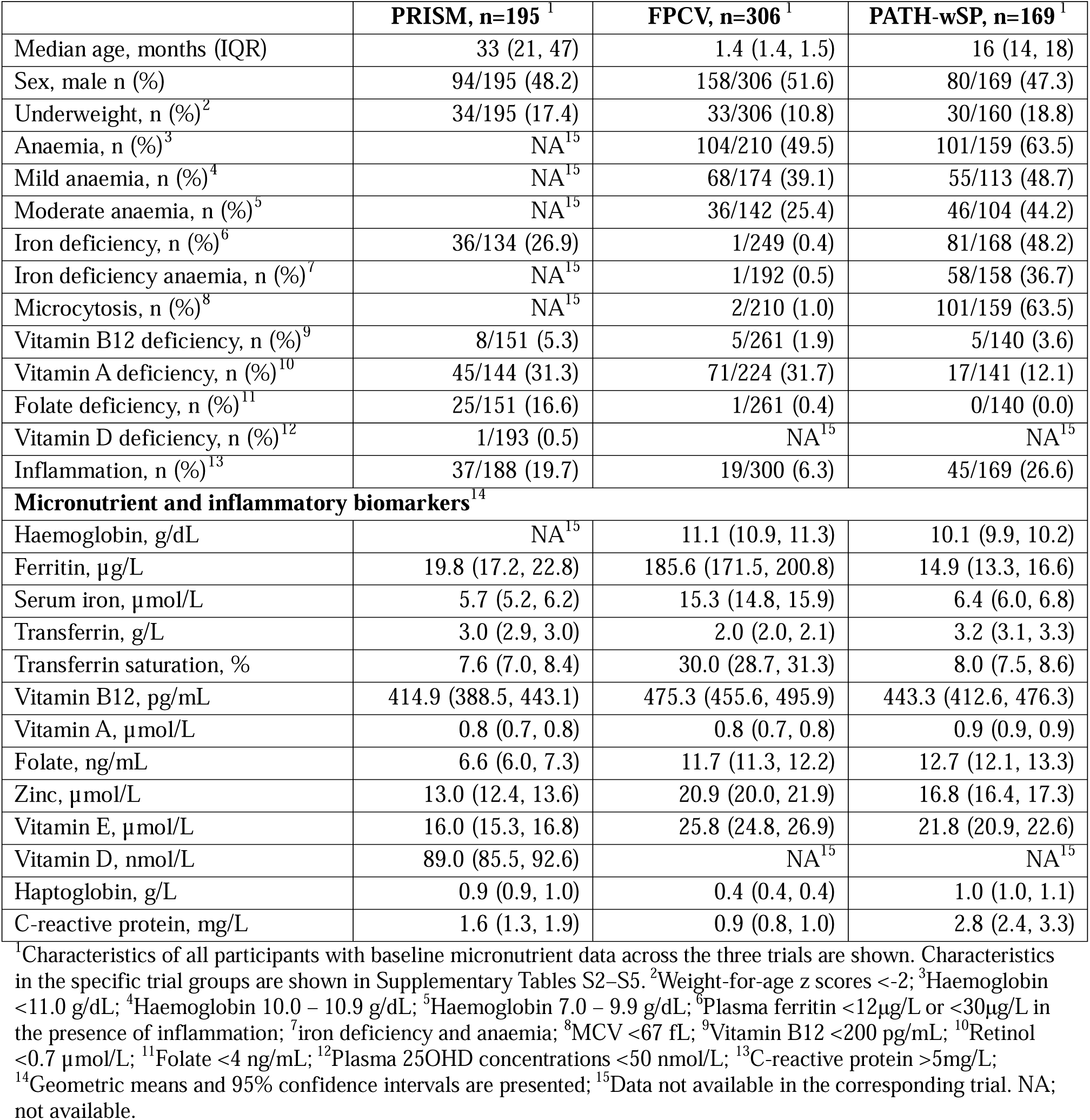
Baseline characteristics of study participants.

Participants in the FPCV trial were younger (median age 1.4 months [IQR 1.4, 1.5]) than those in the PATH-wSP (16 months [14, 18]) and PRISM (33 months [21, 47]) trials. Underweight prevalence was also lower in the FPCV (10.8% [33/306]) than in the PRISM (17.4% [34/195]) or PATH-wSP (18.8% [30/160]) cohorts. Participants in the FPCV trial were more iron replete with higher concentrations of ferritin, serum iron, and transferrin saturation (TSAT), and had lower C-reactive protein (CRP) concentrations compared to the other cohorts, while those in the PRISM trial had lower folate, zinc, and vitamin E concentrations (Table 2). Haemoglobin concentrations were unavailable in the PRISM trial; among the remaining cohorts, anaemia was more common in the PATH-wSP trial than in the FPCV trial (63.5% [101/159] vs. 49.5% [104/210]), with a higher prevalence of moderate anaemia (28.9% [46/159] vs. 17.1% [36/210]). Iron deficiency was also more prevalent in the PATH-wSP trial than in the PRISM or FPCV trials (48.2% [81/168], 26.9% [36/134], and 0.4% [1/249], respectively). The prevalence of vitamin B12 deficiency was low (<5.0%) across all three trials (Table 2).

Within each trial, baseline haemoglobin and micronutrient biomarker concentrations were broadly comparable across study groups (Supplementary Tables S2–S5), with similar post-vaccination IgG and opsonophagocytic activity (OPA) responses between groups receiving pneumococcal vaccines (Supplementary Figures S1–S2).

### Haemoglobin and anaemia

Higher haemoglobin concentrations were strongly associated with higher post-vaccination composite IgG z scores among participants receiving the 1 mg PATH-wSP vaccine (adj. β 0.4 [0.2, 0.6]), but not among those receiving full doses of PCV10 or PCV13 in the FPCV trial (Figure 2A-B). Antigen-specific analyses showed consistently positive effect estimates, with statistically significant associations for L460D, BCH0785, StkP, and PiaA (Supplementary Figure S3).

**Figure 2.**
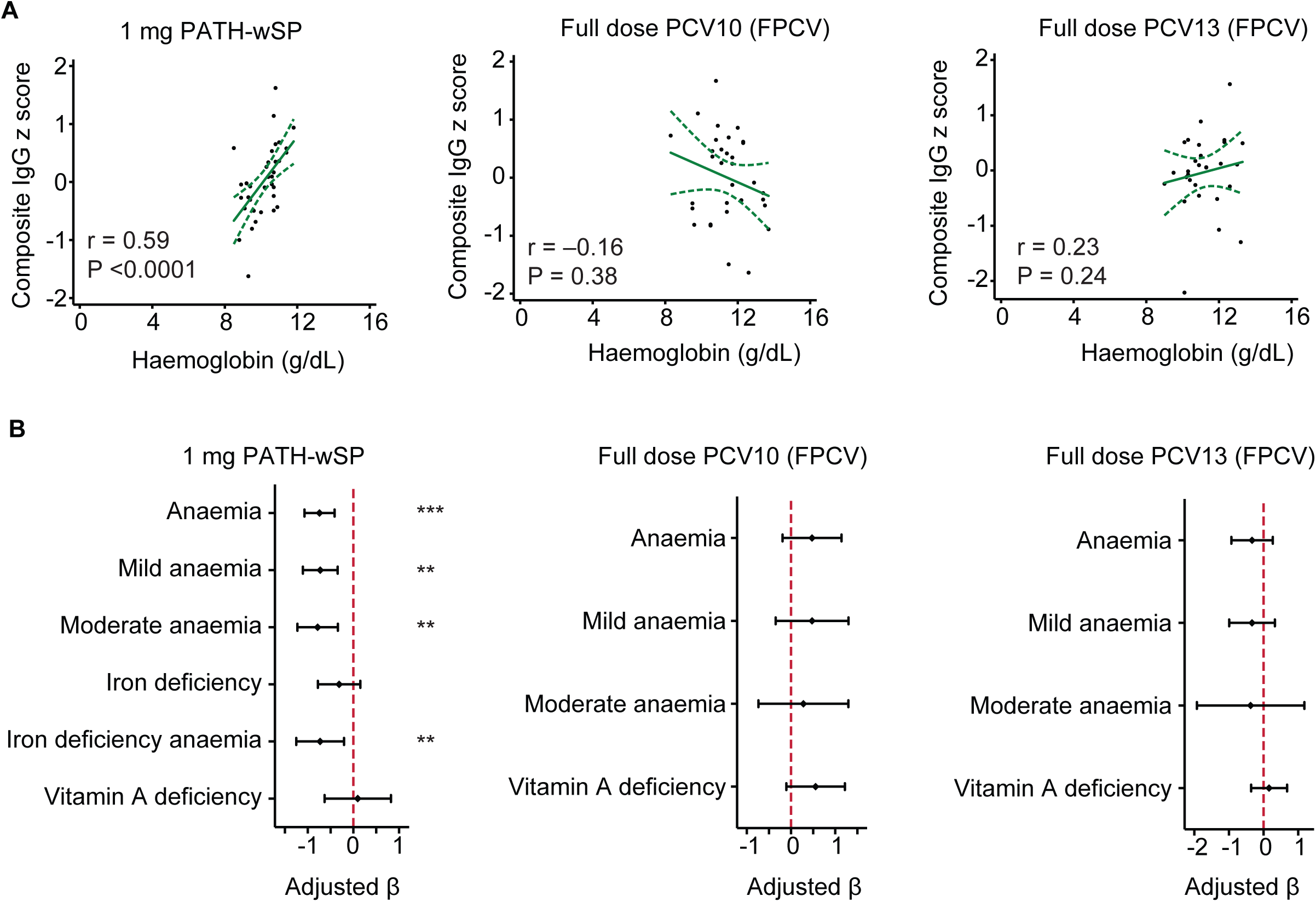
Associations between composite IgG z scores and baseline micronutrient status in the primary full-dose vaccine groups. A) Spearman rank correlations between baseline haemoglobin concentrations and post-vaccination composite IgG z-scores in the primary full-dose vaccine groups. B) Associations between composite IgG z-scores and anaemia, iron deficiency, or vitamin A deficiency in the primary full-dose vaccine groups. Adjusted beta (β) coefficients were estimated using linear regression models adjusted for age, sex, underweight status, and log-transformed C-reactive protein concentrations. Haemoglobin data were not available for the PRISM trial.

Anaemia was also associated with a lower composite IgG z-score (adj. β −0.8 [−1.2, −0.4]), and with lower antigen-specific IgG responses to L460D, BCH0785, StkP, SPWCV, and PiaA among 1 mg PATH-wSP recipients (Supplementary Figure S4-S5). In analyses stratified by anaemia severity, both moderate (adj. β −0.8 [−1.2, −0.3]) and mild anaemia (adj. β −0.8 [−1.3, −0.3]) were associated with lower composite IgG z-scores (Figure 2B). Iron-deficiency anaemia was also associated with a lower composite IgG z score (adj. β −0.7 [−1.2, −0.2]) and lower IgG responses to PhtD, BCH0785, and StkP (Supplementary Figure S5).

Before adjustment for multiple comparisons, moderate anaemia was also associated with lower IgG responses to serotypes 9V (adj. β −2.5 [−4.3, −0.6]) and 14 (adj. β −2.0 [−3.5, −0.4]) among recipients of full PCV13 doses in the FPCV trial (Supplementary Figure S5). These associations were not observed following vaccination with full or fractional PCV10 in the FPCV trial, fractional-dose PCV13, or the 0.6 mg PATH-wSP regimen (Figure 2B, Supplementary Figure S5, Supplementary Table S6).

### Iron Status

Iron deficiency was not associated with composite antibody z-scores in the PRISM or PATH-wSP trials, and few children were iron deficient in the FPCV trial (Supplementary Table S6). Among biomarkers of iron status, ferritin showed a borderline positive association with composite IgG z-scores in the FPCV full-dose PCV13 cohort (adj. β 0.4 [0.0, 0.8]). Before adjustment for multiple comparisons, higher ferritin concentrations were associated with higher IgG responses to serotypes 1 and 6A in the FPCV PCV13 full-dose cohort (Supplementary Figure S6), serotype 7F in the FPCV 40% PCV10 fractional-dose cohort (Supplementary Figure S7), and the StkP antigen in the 1 mg PATH-wSP trial (Supplementary Figure S3); these associations did not persist after Benjamini-Hochberg adjustment. There was little evidence of associations between other biomarkers of iron status and IgG or OPA responses across the three trials (Tables 3-4).

**Table 3.** Associations between baseline micronutrient biomarker concentrations and composite IgG z-scores in the primary full-dose groups across the three vaccine trials.

|  | PCV10<br>(PRISM, n=131)<sup>1</sup> |  | Full dose PCV10<br>(FPCV, n=47)<sup>2</sup> |  | Full-dose PCV13<br>(FPCV, n=42)<sup>2</sup> |  | 1 mg PATH-wSP<br>(n=36)<sup>2</sup> |  |
| --- | --- | --- | --- | --- | --- | --- | --- | --- |
| | Adj. $\beta$ (95% CI)<sup>3</sup> | P<sup>3</sup> | Adj. $\beta$ (95% CI)<sup>3</sup> | P<sup>3</sup> | Adj. $\beta$ (95% CI)<sup>3</sup> | P<sup>3</sup> | Adj. $\beta$ (95% CI)<sup>3</sup> | P<sup>3</sup> |
| Haemoglobin, g/dL | NA<sup>4</sup> | NA<sup>4</sup> | -0.2 (-0.5, 0.0) | 0.07 | 0.1 (-0.2, 0.4) | 0.38 | 0.4 (0.2, 0.6) | 0.0002 |
| Ferritin, μg/L | 0.0 (-0.2, 0.1) | 0.85 | -0.1 (-0.5, 0.3) | 0.69 | 0.4 (0.0, 0.8) | 0.06 | 0.3 (-0.0, 0.6) | 0.10 |
| Serum iron, μmol/L | 0.0 (-0.2, 0.3) | 0.70 | 0.5 (-0.4, 1.4) | 0.24 | 0.7 (-0.1, 1.5) | 0.10 | 0.2 (-0.4, 0.7) | 0.60 |
| Transferrin, g/L | 0.1 (-0.1, 0.3) | 0.55 | 0.4 (-0.2, 1.0) | 0.19 | 0.1 (-0.6, 0.9) | 0.70 | 0.3 (-0.3, 0.9) | 0.28 |
| TSAT, % | 0.0 (-0.2, 0.2) | 0.98 | 0.2 (-0.7, 1.0) | 0.74 | 0.5 (-0.2, 1.1) | 0.14 | 0.1 (-0.5, 0.6) | 0.84 |
| Vitamin B12, pg/mL | 0.4 (0.1, 0.7) | 0.01 | 0.0 (-0.8, 0.8) | 0.94 | 0.3 (-0.4, 0.9) | 0.43 | -0.1 (-0.7, 0.6) | 0.84 |
| Vitamin A, μmol/L | 0.3 (-0.2, 0.7) | 0.22 | 0.4 (-0.8, 1.5) | 0.51 | -0.5 (-1.4, 0.4) | 0.25 | -0.3 (-1.4, 0.8) | 0.56 |
| Folate, ng/mL | -0.1 (-0.3, 0.1) | 0.15 | -0.1 (-1.0, 0.9) | 0.88 | -0.6 (-1.4, 0.2) | 0.16 | 0.2 (-0.7, 1.2) | 0.60 |
| Zinc, μmol/L | 0.0 (-0.4, 0.3) | 0.86 | 0.0 (-0.6, 0.5) | 0.93 | 0.0 (-0.6, 0.7) | 0.93 | 0.3 (-1.0, 1.6) | 0.65 |
| Vitamin E, μmol/L | 0.1 (-0.6, 0.3) | 0.57 | -0.2 (-1.3, 0.9) | 0.70 | -1.1 (-2.1, 0.0) | 0.04 | 0.4 (-0.7, 1.5) | 0.45 |
| Vitamin D, nmol/L | 0.1 (-0.3, 0.5) | 0.81 | NA<sup>4</sup> | NA<sup>4</sup> | NA<sup>4</sup> | NA<sup>4</sup> | NA<sup>4</sup> | NA<sup>4</sup> |

<sup>1</sup>In the PRISM trial, Groups A and B were combined because both received their first PCV10 dose at baseline. <sup>2</sup>Only primary groups receiving full pneumococcal vaccine doses are shown. Fractional-dose vaccine groups, the Kenyan Ministry of Health PCV10 (3p+0) schedule, and the 0.6 mg PATH-wSP group are presented in Supplementary Table S7. <sup>3</sup>Adjusted regression coefficients ( $\beta$ ) and P values were estimated using linear regression models of composite IgG z scores and log-transformed biomarkers (except haemoglobin and transferrin) adjusted for age, sex, underweight status, and log-transformed C-reactive protein concentrations. IgG responses were measured in serum samples collected 30 days after vaccination with one PCV10 dose in the PRISM trial and 28 days after the second vaccine dose in the FPCV and PATH-wSP trials. <sup>4</sup>Biomarker not measured in the corresponding trial. TSAT denotes transferrin saturation; PCV, pneumococcal conjugate vaccine; NA; not available.

**Table 4.** Associations between baseline micronutrient biomarkers and composite opsonophagocytic activity (OPA) z-scores in the PRISM trial.

| | Adj. $\beta$ (95% CI)<sup>1</sup> | P<sup>1</sup> |
| --- | --- | --- |
| Ferritin, $\mu\text{g/L}$ | -0.1 (-0.3, 0.0) | 0.07 |
| Serum iron, $\mu\text{mol/L}$ | 0.0 (-0.2, 0.2) | 0.74 |
| Transferrin, g/L | 0.2 (0.0, 0.3) | 0.05 |
| TSAT, % | 0.0 (-0.2, 0.1) | 0.71 |
| Vitamin B12, pg/mL | 0.4 (0.1, 0.7) | 0.005 |
| Vitamin A, $\mu\text{mol/L}$ | 0.1 (-0.2, 0.5) | 0.45 |
| Folate, ng/mL | -0.1 (-0.3, 0.1) | 0.20 |
| Zinc, $\mu\text{mol/L}$ | 0.1 (-0.2, 0.4) | 0.38 |
| Vitamin E, $\mu\text{mol/L}$ | 0.0 (-0.4, 0.4) | 0.83 |
| Vitamin D, nmol/L | 0.0 (0.0, 0.0) | 0.36 |

**Abbreviations:** <sup>1</sup>Adjusted regression coefficients ( $\beta$ ) and P values were estimated using linear regression models of composite IgG z scores and log-transformed biomarkers (except transferrin) adjusted for age, sex, underweight status, and log-transformed C-reactive protein concentrations. Opsonophagocytic activity was measured in serum samples collected 30 days after vaccination with one PCV10 dose. Haemoglobin concentrations were not available in the PRISM trial.

### Vitamin B12

Higher vitamin B12 concentrations were associated with higher composite IgG z-scores (adj. β 0.4 [0.1, 0.7]) and OPA z-scores (adj. β 0.4 [0.2, 0.7]) in the PRISM trial. Serotype-specific analyses showed consistently positive effect estimates, with statistically significant associations observed for IgG responses to serotypes 1 and 6B, and OPA responses to serotypes 1, 4, and 14 (Figure 3A). Consistent with these findings, vitamin B12 deficiency, present in 4.0% of 101 PCV10-vaccinated children, was associated with lower composite IgG z-scores (adj. β −0.6 [−1.2, −0.3]) and OPA z-scores (adj. β −0.6 [−1.0, −0.1]). Among individual serotypes, vitamin B12 deficiency was associated with lower IgG responses to serotypes 9V and 14 and lower OPA responses to serotypes 4, 9V, and 14 (Figure 3B).

**Figure 3.**
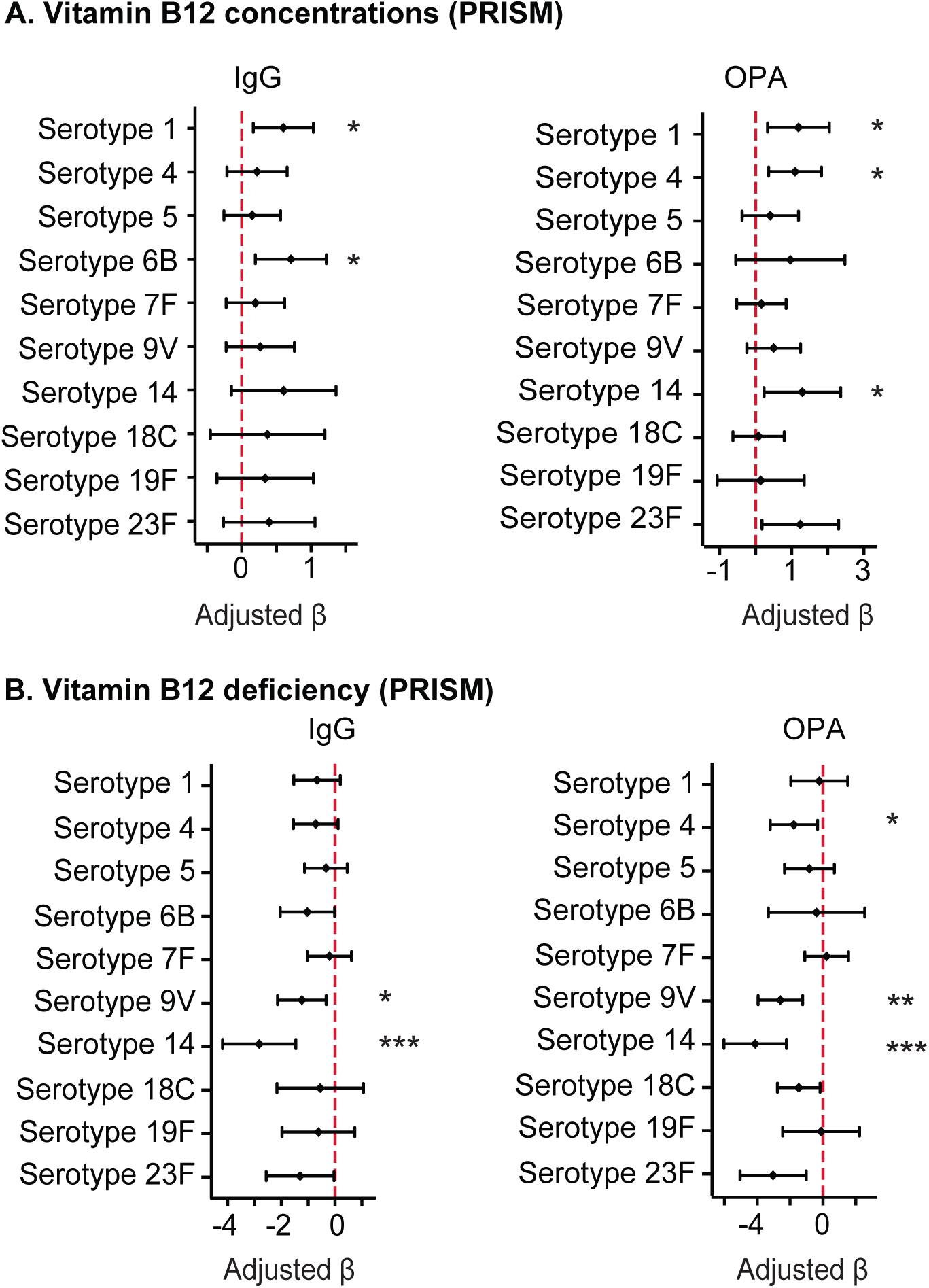
Associations between pneumococcal vaccine antibody responses and vitamin B12 status in the PRISM trial. A) Baseline vitamin B12 concentrations. B) Baseline vitamin B12 deficiency (vitamin B12 <200 pg/mL, n=4/101). Participants in Groups A and B received one dose at baseline and were combined in this analysis. IgG and OPA responses were measured 30 days post vaccination. Adjusted regression coefficients (β) were derived from linear regression models adjusting for age, sex, underweight status, and log-transformed C-reactive protein levels. Statistical significance after Benjamini-Hochberg correction for multiple antibody comparisons: * denotes P<0.05; **, P<0.01; and ***, P<0.001.

In the FPCV and PATH-wSP trials, vitamin B12 concentrations were not associated with composite IgG z-scores (Supplementary Figure S8; Supplementary Tables S7). Before adjustment for multiple comparisons, higher vitamin B12 concentrations were associated with increased IgG responses to serotype 6B following PCV13 vaccination and serotype 9V following PCV10 vaccination in the FPCV trial (Supplementary Figure S8); neither association persisted after Benjamini-Hochberg adjustment.

### Other micronutrient biomarkers

Vitamin A concentrations and deficiency were not associated with pneumococcal vaccine IgG and OPA responses (Figure 2B; Tables 3–4; Supplementary Figure S9). There was little evidence that zinc, folate, vitamin E, or vitamin D concentrations were consistently associated with composite IgG or OPA z-scores or antibody responses to individual serotypes or antigens (Tables 3–4; Supplementary Tables S7–S8; Supplementary Figures S3, S6–S7, S10–S11). In the FPCV trial, higher vitamin E concentrations were associated with borderline lower composite IgG z-scores following full-dose PCV13 (adj. β −1.0 [−2.0, 0.0]), fractional 20% PCV13 (adj. β −1.3 [−2.6, −0.1]), and PCV10 administered according to the Kenyan Ministry of Health schedule (adj. β −0.9 [−1.8, −0.1]) (Supplementary Table S7; Supplementary Figure S12). Higher folate concentrations were also associated with lower composite IgG z-scores following fractional 20% PCV13 vaccination (adj. β −0.8 [−1.6, −0.1]).

## Discussion

Across three pneumococcal vaccine trials in Kenyan children, anaemia and vitamin B12 concentrations were associated with post-vaccination antibody responses in specific vaccine and age contexts. There was little evidence of associations for most other micronutrients. In the PATH-wSP trial, anaemia was associated with lower antibody responses following the 1 mg dose, with higher haemoglobin concentrations associated with stronger responses. Higher vitamin B12 concentrations were positively associated with post-PCV10 vaccination composite IgG and OPA responses in the PRISM trial, with consistent associations across multiple individual serotypes. In contrast, there was little evidence that ferritin, vitamin A, folate, zinc, vitamin E, or vitamin D concentrations were associated with pneumococcal vaccine-induced antibody responses. Overall, these findings suggest that the relationship between micronutrient status and pneumococcal vaccine immunogenicity is context-dependent, and varies by micronutrient, age, and vaccine platform.

In the PATH-wSP trial, anaemia was associated with weaker pneumococcal vaccine responses, while higher haemoglobin concentrations were associated with stronger responses following the 1 mg dose. Mild, moderate, and iron-deficiency anaemia were associated with lower composite and antigen-specific responses. These findings are consistent with evidence linking anaemia to impaired vaccine responses,^14,17–19^ and with increased risk of invasive bacterial infection among children with moderate or severe anaemia.^11,20^ In contrast, we found little evidence that iron deficiency or individual biomarkers of iron status were consistently associated with vaccine antibody responses. Ferritin concentrations showed a borderline positive association with PCV13 responses in the FPCV trial, but no consistent associations were observed for other iron biomarkers or across cohorts. This contrasts with previous observations from south coastal Kenya, where iron deficiency was associated with antibody responses to pneumococcal serotype 19, although iron status in that study was assessed using soluble transferrin receptor concentrations.^14^ The apparent disconnect between anaemia and conventional measures of iron status raises several possible explanations. First, iron deficiency may need to be sufficiently severe to result in anaemia before affecting vaccine responses. Anaemia itself may also influence immunogenicity through pathways beyond iron deficiency, including tissue hypoxia and altered cellular proliferation.^21–23^ Ferritin concentrations were also high, particularly among the 6-week old infants in the FPCV trial, potentially reflecting the distinct iron physiology of early infancy.^24^ Consequently, developmental changes in iron homeostasis, together with residual inflammation, may have obscured associations between iron status and vaccine responses.

Vitamin B12 concentrations were positively associated with composite IgG and OPA responses and showed broadly consistent positive associations across individual PCV10 serotypes in the PRISM trial. This finding is consistent with a previous study of institutionalised older adults, where vitamin B12 deficiency impaired antibody responses to 12 serotypes of the 23-valent pneumococcal polysaccharide vaccine.^15^ Although vitamin B12 deficiency was uncommon in the PRISM cohort, lower concentrations were associated with reduced antibody responses, suggesting that variation in B12 status within the non-deficient range may also be associated with pneumococcal vaccine immunogenicity. Vitamin B12 is essential for DNA synthesis, cellular proliferation, and one-carbon metabolism,^25,26^ processes that support clonal expansion of antigen-specific B cells, germinal centre formation, and development of effective T cell responses.^12,26,27^ Positive effect estimates across most serotypes supported the composite analyses, although statistically significant associations were confined to relatively fewer immunogenic serotypes, including serotypes 1 and 6B. One possibility is that nutritional constraints become more apparent when vaccine-induced responses are intrinsically weaker, as has been reported for serotype 6B.^28,29^ However, this association was not observed in the PATH-wSP or FPCV cohorts, suggesting that the associations of vitamin B12 status and pneumococcal vaccine responses may depend on various factors including age, vaccine platform, or nutritional context rather than representing a uniform effect across populations. Larger, adequately powered studies of vitamin B12 deficiency are needed to replicate these findings.

We found limited evidence that concentrations of vitamin A, folate, zinc, vitamin E, or vitamin D were associated with pneumococcal vaccine responses. These findings are broadly consistent with the limited and heterogeneous literature on micronutrient status and pneumococcal vaccine responses. Vitamin A supplementation did not enhance pneumococcal vaccine responses in retinol-sufficient children or in people living with HIV who inject drugs.^30,31^ Zinc supplementation similarly did not enhance pneumococcal conjugate vaccine responses people living with HIV who inject drugs or in patients with colorectal cancer,^31,32^ whereas it improved responses to serotype 9V in Bangladeshi infants^33^. Vitamin D deficiency was associated with impaired pneumococcal vaccine responses in patients with asthma, but this association was not observed in healthy controls.^34^ Folate status was not associated with antibody responses to a 23-valent pneumococcal vaccine among institutionalised older adults,^15^ consistent with the limited evidence for folate–vaccine response associations in our study, although we observed nominal associations with PCV13 responses following fractional 20% dosing. We found that higher vitamin E concentrations were associated with lower composite IgG z-scores in some FPCV groups, contrasting with evidence that vitamin E supplementation can enhance pneumococcal vaccine responses in healthy older adults.^35^ These contrasting findings may reflect differences in nutritional status, age, vaccine platform, or study population. The low prevalence of severe micronutrient deficiencies in our cohorts may have limited our ability to detect effects occurring primarily in the extremes of nutritional status. Our findings, therefore, do not exclude effects in children with more severe deficiencies or in populations with greater nutritional vulnerability. In addition, immune pathways regulated by micronutrients are highly interconnected,^12^ which potentially allows for compensation when variation in a single micronutrient is modest.

A strength of this study is the evaluation of multiple micronutrient biomarkers across three pneumococcal vaccine trials, enabling comparisons across vaccine platforms, dosing strategies, age groups, and nutritional biomarkers. However, some limitations should be considered. First, the observational design precludes causal inference. Residual confounding by unmeasured factors, including diet, breastfeeding, maternal antibodies, genetic variation, and environmental exposures, cannot be excluded. Second, several subgroup analyses involved relatively small numbers of participants, and the low prevalence of micronutrient deficiencies limited statistical power to detect associations concentrated at the extremes of nutritional status. Although composite z scores provide a summary measure of overall vaccine response and reduce the dimensionality of multiple serotype-specific outcomes, they may be less sensitive to biologically meaningful effects confined to a subset of serotypes.

Third, the large number of biomarker, serotype, and antigen comparisons increased the potential for type I error. We controlled the false discovery rate using the Benjamini– Hochberg method, and interpreted findings by prioritising composite antibody z-scores, consistent effect estimates across serotypes, and biological plausibility. Fourth, biomarker availability differed between trials: haemoglobin was not measured in PRISM, while vitamin D was not measured in FPCV or PATH-wSP, precluding direct comparison of these biomarkers across all cohorts. Finally, the trials differed in age, vaccine platform, dosing schedule, and eligibility criteria, limiting direct comparison of effect estimates across cohorts despite separate within-trial analyses. All three trials were conducted in coastal Kenya and excluded children with severe anaemia, severe malnutrition, and sickle cell disease, which may limit generalisability to populations with greater nutritional vulnerability or different epidemiological and healthcare contexts.

In conclusion, anaemia and vitamin B12 concentrations were associated with context-dependent pneumococcal vaccine immunogenicity. Evidence for associations with other micronutrient biomarkers was limited. These findings suggest that nutritional status is not a uniform determinant of pneumococcal vaccine immunogenicity, but that specific nutritional factors may be associated with vaccine responses in particular age, vaccine, and dosing contexts. Larger cohorts are needed to replicate these findings and determine their generalisability across populations and vaccine platforms. If confirmed, prospective intervention studies should assess whether correcting anaemia or vitamin B12 deficiency can improve vaccine responses, particularly in populations with a high burden of nutritional deficiencies. Such strategies may become increasingly important as higher-valency PCVs, including PCV20, are introduced, given that increasing serotype valency may be accompanied by lower serotype-specific immunogenicity.^36^

## Methods

### Study design and participants

This retrospective observational study analysed data and archived serum samples from children living in Kilifi, Kenya, who were enrolled in three randomised clinical trials evaluating pneumococcal vaccines. We investigated associations between baseline anaemia or micronutrient status, and humoral immune responses following pneumococcal vaccination. A total of 670 children with immunogenicity data, sufficient archived sample for micronutrient biomarker measurements, and prior consent for future research use of samples were included. Micronutrient biomarkers and haemoglobin concentrations were measured in baseline, pre-vaccination samples.

### PRISM (PCV10 Reactogenicity and Immunogenicity Study - Malindi, NCT01028326)

This phase III trial assessed the reactogenicity and immunogenicity of the 10-valent pneumococcal non-typeable *Haemophilus influenzae* protein-D conjugate vaccine (PCV10, GlaxoSmithKline plc. [GSK]).^37^ Conducted between January and September 2010, 600 children aged 12–59 months were randomised equally to receive two doses of PCV10 60 days apart followed by diphtheria, tetanus, and acellular pertussis (DTaP) vaccine at day 180 (Group A), two doses of PCV10 180 days apart with DTaP administered at day 60 (Group B), or two doses of hepatitis A vaccine (HAV) 180 days apart with DTaP administered at day 60 (Group C, control group). Blood samples were collected at baseline, 30 days after the first vaccination, and 30 days after either the second (Groups A and C) or third vaccination (Group B). Serotype-specific immune responses were measured in an immunogenicity cohort of 375 participants. For the present study, 195 children with sufficient archived serum for micronutrient biomarker measurements were included (Figure 1). Haemoglobin measurements were not available in the PRISM study.

### FPCV (Fractional Doses of Pneumococcal Conjugate Vaccine, NCT03489018)

Conducted between March 2019 and November 2021, the trial enrolled healthy infants aged 6–8 weeks who were randomised to receive either full or fractional doses of PCV10 (GSK) or PCV13 (Pfizer).^38^ Participants in Groups A–F received a two-dose primary series at 6 and 14 weeks followed by a booster dose at 9 months (2p+1 schedule), while a comparator group (Group G) received full-dose PCV10 according to the Kenyan Ministry of Health Expanded Programme on Immunization schedule (3p+0, Figure 1). Blood samples were collected at baseline, 4 weeks after completion of the primary doses, and 4 weeks after the booster dose. Baseline micronutrient status was assessed in 306 participants with available archived serum samples. Analyses were restricted to immune responses following the primary series because baseline micronutrient status was unlikely to reflect nutritional status at the time of booster vaccination 8 months later.

### PATH-wSP (Inactivated Streptococcus pneumoniae Whole-Cell Vaccine, NCT02543892)

This phase I/II randomised, placebo-controlled, quadruple-blind trial evaluated the safety, tolerability, and immunogenicity of the PATH-wSP vaccine, an inactivated whole-cell *S. pneumoniae* vaccine adsorbed to aluminium hydroxide (alum). Conducted between August 2016 and January 2018, the trial enrolled young Kenyan adults (n=48) and toddlers aged 12– 21 months (n=200). Only toddlers with sufficient baseline sera for micronutrient biomarker measurement (n=169) were included in the present study (Figure 1). Participants were randomised to receive two doses of either 0.6 mg or 1 mg of the PATH-wSP vaccine or placebo (saline), administered 28 days apart. The second dose was co-administered with a pentavalent vaccine (diphtheria, tetanus, whole-cell pertussis, *Haemophilus iinfluenzae* type b, and hepatitis B). Immunogenicity was assessed at baseline and 4 weeks after the second vaccination. The PATH-wSP vaccine candidate has not progressed beyond Phase II clinical development.

### Procedures

Vaccine-type capsular polysaccharide-specific IgG responses were quantified using ELISA in the PRISM and FPCV trials.^37,38^ IgG responses were measured against all vaccine serotypes, except in the Kenyan Ministry of Health schedule group (Group G), in which responses were measured to seven PCV10 serotypes.^38^ In the PATH-wSP trial, IgG responses to nine pre-specified pneumococcal protein antigens were quantified using the Meso Scale Discovery electrochemiluminescence multiplex platform (Supplementary Table S1).

Functional antibody responses in the PRISM trial were assessed by OPA using a modified WHO HL-60 reference assay at the SGS laboratory in Belgium.^39^ Viable colony counts were determined after overnight incubation of plates at 37°C in a 5% CO₂ atmosphere. OPA titres were defined as the reciprocal serum dilution achieving ≥50% bacterial killing. Among PCV recipients in the PRISM and FPCV trials, vaccine protection was defined as serotype-specific IgG concentrations ≥0.35 µg/mL or OPA titre ≥8.^37,38^

Haemoglobin concentrations (FPCV and PATH-wSP) were measured using a Beckman Coulter haematology analyser (Brea, CA, USA). Baseline micronutrient biomarkers were measured in serum samples stored at −80°C. Ferritin, serum iron, transferrin, haptoglobin, zinc, and CRP were quantified using a Siemens Dimension Xpand clinical chemistry analyser (Siemens Healthcare Ltd, Camberley, Surrey, UK). Vitamin B12 and folate concentrations were measured using the Tosoh AIA-CL1200 immunoassay analyser (Tosoh Bioscience, Tokyo, Japan) according to the manufacturer’s instructions. In the PRISM trial, 25-hydroxyvitamin D [25(OH)D] concentrations were measured using the VIDAS® enzyme-linked fluorescent assay (ELFA; bioMérieux, Marcy-l’Étoile, France). Vitamin A (retinol) and vitamin E (α- and γ-tocopherol) concentrations were quantified by liquid–liquid extraction followed by high-performance liquid chromatography (HPLC) using a YMC-Pack Pro C18 analytical column. Concentrations were calculated from calibration curves based on the ratio of analyte-to-internal standard peak area. Although both α- and γ-tocopherol were quantified, only α-tocopherol was included in regression analyses because it is the principal circulating biomarker of vitamin E status.^40^

### Definitions

Anaemia was defined according to age-specific thresholds from the World Health Organization^41^ and the Global Burden of Disease Anaemia Collaborators^8^ as haemoglobin (Hb) <11.0 g/dL in children aged 1–5 months and 24–59 months, or Hb <10.5 g/dL in those aged 6–23 months. Mild anaemia was defined as Hb 10.0–10.9 g/dL (1–5 and 24–59 months) or 9.5–10.4 g/dL (6–23 months); moderate anaemia as Hb 7.0–9.9 g/dL (1–5 and 24–59 months) or 7.0–9.4 g/dL (6–23 months); and severe anaemia as Hb <7.0 g/dL (1–59 months). Inflammation was defined as CRP concentration ≥5 mg/l; iron deficiency as ferritin <12 µg/L, or ferritin <30 µg/L in children with inflammation^42^; iron deficiency anaemia as the concurrent presence of iron deficiency and anaemia; vitamin B12 deficiency as plasma vitamin B12 <200 pg/mL^43^; folate deficiency as serum folate <4 ng/mL^44^; vitamin A deficiency as plasma retinol <0.7 µmol/L^45^; vitamin D deficiency as 25(OH)D <50 nmol/L^46^; and underweight as weight-for-age z scores <-2^47^. TSAT was calculated as 100 × serum iron (µmol/L)/(transferrin (g/L) × 25.1).

### Outcomes

The primary outcome was the post-vaccination composite antibody response z-score. Secondary outcomes were serotype- or antigen-specific IgG concentrations and OPA titres. Outcomes were assessed 30 days after a single PCV10 dose in the PRISM trial, 28 days after primary vaccination in the FPCV trial, and 28 days after the second PATH-wSP dose. In PRISM, Groups A and B both received their first PCV10 dose on day 0; therefore, analyses of antibody responses following the first dose combined these groups.

### Statistical analysis

Statistical analyses were performed with Stata 15.1 (StataCorp., College Station, TX). Because the trials differed in age groups, vaccine platforms, and immunisation schedules (Table 1), analyses were conducted separately by trial and, where appropriate, by vaccine group. To provide a common measure of overall vaccine immunogenicity, log-transformed serotype- or antigen-specific antibody responses were standardised to z scores within each trial and vaccine group using the formula: *z* = (x − μ)/σ, where *x* is the individual log-transformed antibody response, *μ* is the mean response, and *σ* is the group standard deviation for that serotype or antigen.^48^ For each participant, the composite antibody response z-score was calculated as the mean of the standardised z-scores across all measured serotypes or antigens.^49^

Categorical variables were summarised as frequencies with percentages and compared using Fisher’s exact or Chi-square tests, as appropriate. Continuous variables were summarised as geometric means with 95% confidence intervals (CI) or medians with interquartile ranges (IQR). Distributional assumptions were assessed using histograms and the Shapiro–Wilk test. Skewed micronutrient biomarkers (except haemoglobin and transferrin, which were approximately normally distributed), IgG concentrations, and OPA titres were natural log transformed.

Univariable and multivariable least-squares linear regression models were used to assess associations between micronutrient status and antibody responses. Primary analyses evaluated continuous micronutrient biomarker concentrations in relation to composite antibody z-scores. Secondary analyses evaluated individual post-vaccination log-transformed IgG concentrations and OPA titres, as well as clinically defined categories of anaemia and micronutrient deficiency. Covariates were prespecified based on biological plausibility and evidence from the literature and included age, sex, underweight status, and log-transformed CRP concentration. Linear regression effect estimates are reported as adjusted β coefficients with 95% CIs. For each biomarker, adjusted P-values for associations with individual serotypes were calculated using the Benjamini–Hochberg method to control the false discovery rate at 5%.^50^ Regression analyses were performed using complete-case data, and missing data were assumed to be missing at random. Spearman’s rank correlation was used to assess correlations between micronutrient biomarkers and composite antibody z scores. All P-values are two-sided.

### Ethics statement

Ethical approval for the analysis of stored samples and data from the vaccine trials was granted by the Kenya Medical Research Institute (KEMRI) Scientific and Ethics Review Unit (KEMRI/SERU/CGMR-C/269/4545). Written informed consent was obtained from the parents or legal guardians of all participants in the original vaccine trials. The study is reported in accordance with STROBE guidelines.

## Acknowledgements

We thank all study participants who contributed to this study, the VAnguard team, and staff involved with consent, sample and data collection and preparation. This manuscript was submitted for publication with the permission of the Director of the Kenya Medical Research Institute (KEMRI).

## Author contributions

Conceptualisation: KMA, HKK, AME, SHA. Investigation: KMA, HKK, KG, BWM, BM, AK, EMI, JMM, LH, SK, CAM, MN, JAGS, AME, GN, SHA. Formal analysis: KMA, HKK, CKK, SHA. Supervision: SK, CAM, MN, JAGS, AME, GN, SHA. Writing – original draft: KMA, SHA. Writing – review and editing: KMA, HKK, KG, BWM, CKK, BM, AK, EMI, JMM, LH, SK, CAM, MN, JAGS, AME, GN, SHA. All authors had full access to the data. KMA, HK, CKK, and SHA accessed and verified the data. No authors were prohibited from accessing the data. All authors critically reviewed and approved the manuscript and had final responsibility for the decision to submit for publication.

## Data availability statement

All data and analyses underlying this article are available in Harvard Dataverse repository (https://doi.org/10.7910/DVN/J0L1SO) and applications for data access can be made through the Kilifi Data Governance Committee.

