## Supplementary File for "Association between anaemia, micronutrient status, and pneumococcal vaccine responses in young Kenyan children"

^1^KEMRI-Wellcome Trust Research Programme, Kilifi, Kenya; ^2^Immunomodulation and Vaccines Group, Vaccine Research Theme, Medical Research Council/Uganda Virus Research Institute and London School of Hygiene and Tropical Medicine (MRC/UVRI and LSHTM) Uganda Research Unit, Entebbe, Uganda; ^3^Department of Infectious Disease Epidemiology, London School of Hygiene and Tropical Medicine, London, UK; ^4^Department of Infection Biology, London School of Hygiene and Tropical Medicine, London, UK; ^5^Department of International Health, Johns Hopkins Bloomberg School of Public Health, Baltimore, MD, USA; ^6^Department of Infectious Diseases, Imperial College London, London, UK; ^7^Department of Immunology and Immunotherapy, College of Medical and Dental Sciences, University of Birmingham, Birmingham, UK; ^8^Department of Internal Medicine II, Medical University Innsbruck, Innsbruck, Austria; ^9^Department of Clinical Research, London School of Hygiene and Tropical Medicine, London, UK; ^10^Centre for Tropical Medicine and Global Health, Nuffield Department of Clinical Medicine, University of Oxford, Oxford, UK; and ^11^Department of Paediatrics, University of Oxford, Oxford, UK.

**Supplementary Tables**

**Table S1.** Pneumococcal proteins assessed in the PATH-wSP vaccine trial

| **Protein** | **Location** | **Function** |
| --- | --- | --- |
| L460D | Cytoplasmic | Metabolic processes and bacterial stress response. |
| PspA-Fam1 | Cell surface | Complement inhibition and immune evasion. |
| PhtD | Cell surface | Adhesion, virulence, complement inhibition. |
| BCH0785 | Cytoplasmic | Bacterial metabolism (function unclear). |
| StkP | Membrane-associated | Cell division and stress responses. |
| PcpA | Cell surface | Adherence and colonisation, choline binding protein. |
| SPWCV | Whole cell | Whole cell antigens (function unclear). |
| PiuA | Cell surface | Iron acquisition through haem uptake (ABC transporter). |
| PiaA | Cell surface | Iron uptake (ABC transporter). |

**Abbreviations**: PspA-Fam1 denotes pneumococcal surface protein A family 1; PhtD, Pneumococcal histidine triad protein D; BCH0785, Bacterial Conserved Hypothetical Protein 0785; StkP, serine/threonine kinase protein; PcpA, pneumococcal choline-binding protein A; SPWCV, *Streptococcus pneumoniae* whole cell vaccine antigens; PiuA, pneumococcal iron uptake A; PiaA, pneumococcal iron acquisition A; ABC, ATP-binding cassette.

**Table S2.** Baseline characteristics of participants in the PRISM vaccine trial

|  | **Group C,** ^1^  n=64 | **Group A,** ^1^  n=65 | **Group B,** ^1^  n=66 |
| --- | --- | --- | --- |
| Median age, months (IQR) | 33.0 (19.0, 47.0) | 35.5 (21.5, 46.5) | 31.5 (22.0, 46.0) |
| Sex, male n (%) | 35/64 (54.7) | 26/65 (40.0) | 33/66 (50.0) |
| Underweight, n (%)^2^ | 9/64 (14.1) | 15/65 (23.1) | 10/66 (15.2) |
| Iron deficiency, n (%)^3^ | 15/43 (34.9) | 9/46 (19.6) | 12/45 (26.7) |
| Vitamin B12 deficiency, n (%)^4^ | 4/50 (8.0) | 4/48 (8.3) | 0/53 (0) |
| Vitamin A deficiency, n (%)^5^ | 12/49 (24.5) | 13/44 (29.5) | 20/51 (39.2) |
| Folate deficiency, n (%)^6^ | 9/50 (18.0) | 7/48 (14.6) | 9/53 (17.0) |
| Vitamin D deficiency, n (%)^7^ | 1/64 (1.6) | 0/63 (0) | 0/66 (0) |
| Inflammation, n (%)^8^ | 12/61 (19.7) | 12/63 (19.1) | 13/64 (20.3) |
| **Micronutrient and inflammation biomarkers**^9^ | | | |
| Ferritin, µg/L (n=134) | 16.3 (13.2, 20.0) | 22.9 (17.6, 29.8) | 20.7 (15.8, 27.0) |
| Serum iron, µmol/L (n=167) | 5.7 (4.9, 6.7) | 5.9 (5.1, 6.8) | 5.5 (4.8, 6.3) |
| Transferrin, g/L (n=193) | 3.0 (2.8, 3.1) | 3.0 (2.9, 3.2) | 2.9 (2.8, 3.1) |
| TSAT, % (n=166) | 7.7 (6.5, 9.0) | 7.8 (6.6, 9.3) | 7.4 (6.3, 8.8) |
| Vitamin B12, pg/mL (n=151) | 374.5 (331.1, 423.7) | 434.5 (384.4, 491.1) | 438.3 (396.9, 484.0) |
| Vitamin A, µmol/L (n=144) | 0.8 (0.7, 0.9) | 0.8 (0.7, 0.9) | 0.7 (0.7, 0.8) |
| Folate, ng/mL (n=151) | 6.6 (5.6, 7.8) | 6.9 (5.9, 8.1) | 6.4 (5.3, 7.7) |
| Zinc, µmol/L (n=175) | 12.4 (11.4, 13.6) | 13.3 (12.2, 14.6) | 13.2 (12.2, 14.4) |
| α-tocopherol, µmol/L (n=144) | 16.1 (14.9, 17.3) | 16.3 (14.8, 17.9) | 15.7 (14.5, 17.1) |
| γ-tocopherol, µmol/L (n=144) | 0.9 (0.7, 1.0) | 1.1 (0.9, 1.3) | 0.9 (0.7, 1.0) |
| Vitamin D, nmol/L (n=194) | 88.8 (82.4, 95.6) | 90.1 (84.6, 96.0) | 88.2 (82.0, 94.0) |
| Haptoglobin, g/L (n=192) | 1.0 (0.8, 1.1) | 0.9 (0.8, 1.0) | 0.9 (0.8, 1.1) |
| CRP, mg/L (n=188) | 1.6 (1.2, 2.1) | 1.5 (1.1, 2.0) | 1.7 (1.3, 2.4) |

**Abbreviations**: TSAT denotes transferrin saturation; IQR, interquartile range; and CRP, C-reactive protein. ^1^Participants were assigned into three groups depending on the vaccine sequence on days 0, 60 and 180: Group A received PCV10, PCV10 and DTaP; Group B received PCV10, DTaP, and PCV10; Group C (control group) received hepatitis A vaccine (HAV), DTaP and HAV. Only participants with measured micronutrient biomarkers are included in this analysis. Haemoglobin concentrations were not available in the PRISM trial. ^2^Weight-for-age z scores <-2; ^3^Plasma ferritin <12 µg/L or <30 µg/L in the presence of inflammation; ^4^Vitamin B12 <200 pg/mL; ^5^Retinol <0.7 µmol/L; ^6^Folate <4 ng/mL; ^7^Plasma 25OHD concentrations <50 nmol/L; ^8^C-reactive protein >5mg/L; ^9^Geometric means and 95% confidence intervals are presented.

**Table S3.** Baseline characteristics of participants in the PCV10 groups of the FPCV trial

|  | **Full dose PCV10,** ^1^  n=47 | **40% dose PCV10,** ^1^  n=45 | **20% dose PCV10,** ^1^  n=44 | **MoH Schedule,** ^1^ n=45 |
| --- | --- | --- | --- | --- |
| Median age, months (IQR) | 1.5 (1.4, 1.6) | 1.4 (1.4. 1.5) | 1.4 (1.4, 1.5) | 1.4 (1.4, 1.5) |
| Sex, male n (%) | 22/47 (46.9) | 23/45 (51.1) | 27/44 (61.4) | 18/45 (40.0) |
| Underweight, n (%)^2^ | 6/47 (12.8) | 4/45 (8.9) | 3/44 (6.8) | 7/45 (15.6) |
| Anaemia, n (%)^3^ | 12/32 (37.5) | 19/30 (63.3) | 14/31 (45.2) | 17/35 (48.6) |
| Mild anaemia, n (%)^4^ | 7/32 (21.9) | 13/30 (43.3) | 10/31 (32.3) | 9/35 (25.7) |
| Moderate anaemia, n (%)^5^ | 5/32 (15.6) | 6/30 (20.0) | 4/31 (12.9) | 8/35 (22.9) |
| Microcytosis, n (%)^6^ | 0/32 (0) | 0/30 (0) | 0/31 (0) | 1/35 (2.9) |
| Vitamin B12 deficiency, n (%)^7^ | 0/41 (0) | 2/36 (5.6) | 0/37 (0) | 0/39 (0) |
| Vitamin A deficiency, n (%)^8^ | 10/35 (28.6) | 8/23 (34.8) | 11/36 (30.6) | 16/36 (44.4) |
| Folate deficiency, n (%)^9^ | 0/41 (0) | 0/36 (0) | 0/37 (0) | 1/39 (2.6) |
| Inflammation, n (%)^10^ | 2/46 (4.4) | 2/43 (4.7) | 4/44 (9.1) | 2/44 (4.6) |
| **Micronutrient and inflammation biomarkers**^11^ | | | |  |
| Haemoglobin (n=116), g/dL | 11.0 (10.6, 11.4) | 11.0 (10.6, 11.5) | 10.9 (10.4, 11.4) | 10.9 (10.4, 11.4) |
| Ferritin (n=111), µg/L | 187.8 (153.4, 230.0) | 165.9 (139.4, 197.4) | 211.3 (171.7, 260.0) | 213.6 (174.2, 261.8) |
| Serum iron (n=118), µmol/L | 15.8 (14.5, 17.2) | 14.5 (13.0, 16.3) | 15.5 (14.2, 17.1) | 15.6 (13.9, 17.6) |
| Transferrin (n=131), g/L | 2.0 (1.9, 2.2) | 2.0 (1.9, 2.1) | 2.0 (1.8, 2.1) | 2.0 (1.9, 2.2) |
| TSAT (n=118), % | 30.5 (27.9, 33.3) | 28.8 (25.3, 32.8) | 31.8 (28.0, 36.0) | 30.0 (25.9, 34.8) |
| MCV (n=93), fL | 85.6 (82.9, 88.3) | 87.3 (84.8, 89.9) | 88.1 (85.3, 90.9) | 85.8 (82.9, 88.9) |
| Vitamin B12 (n=114), pg/mL | 471.4 (424.9, 523.1) | 466.9 (412.3, 528.7) | 520.0 (472.0, 572.7) | 484.8 (437.6, 537.1) |
| Vitamin A (n=94), µmol/L | 0.8 (0.7, 0.9) | 0.8 (0.7, 0.9) | 0.7 (0.7, 0.8) | 0.7 (0.7, 0.8) |
| Folate (n=114), ng/mL | 12.0 (11.0, 13.0) | 11.6 (10.4, 12.9) | 11.9 (10.9, 12.9) | 10.9 (9.6, 12.4) |
| Zinc (n=122), µmol/L | 21.8 (18.9, 25.2) | 22.4 (20.1, 25.0) | 20.8 (18.4, 23.5) | 20.1 (17.8, 22.6) |
| α-tocopherol (n=94), µmol/L | 26.2 (24.0, 28.6) | 28.9 (26.2, 32.0) | 26.6 (24.0, 29.6) | 25.2 (22.6, 28.0) |
| γ-tocopherol (n=94), µmol/L | 0.9 (0.8, 1.0) | 0.9 (0.7, 1.2) | 1.0 (0.8, 1.1) | 1.0 (0.8, 1.2) |
| Haptoglobin (n=131), g/L | 0.4 (0.3, 0.6) | 0.4 (0.3, 0.5) | 0.4 (0.3, 0.5) | 0.5 (0.3, 0.6) |
| CRP (n=133), mg/L | 0.8 (0.6, 1.1) | 0.9 (0.6, 1.2) | 0.9 (0.6, 1.2) | 1.0 (0.7, 1.3) |

**Abbreviations**: TSAT denotes transferrin saturation; MCV, mean corpuscular volume; IQR, interquartile range; and CRP, C-reactive protein. ^1^Only participants with measured micronutrient biomarkers are included in this analysis. Iron deficiency was not observed in this cohort. ^2^Weight-for-age z scores <-2; ^3^Haemoglobin <11.0 g/dL; ^4^Haemoglobin 10.0 – 10.9 g/dL; ^5^Haemoglobin 7.0 – 9.9 g/dL; ^6^MCV <67 fL; ^7^Vitamin B12 <200 pg/mL; ^8^Retinol <0.7 µmol/L; ^9^Folate <4 ng/mL; ^10^C-reactive protein >5mg/L; ^11^Geometric means and 95% confidence intervals are presented.

**Table S****4.** Baseline characteristics of participants in the PCV13 groups of the FPCV trial

|  | **Full dose PCV13,** ^1^  n=42 | **40% dose PCV13,** ^1^  n=39 | **20% dose PCV13,** ^1^  n=44 |
| --- | --- | --- | --- |
| Median age, months (IQR) | 1.41 (1.38, 1.51) | 1.41 (1.38. 1.48) | 1.46 (1.40, 1.54) |
| Sex, male n (%) | 24/42 (57.1) | 21/39 (53.4) | 23/44 (52.3) |
| Underweight, n (%)^2^ | 5/42 (11.9) | 5/39 (12.8) | 3/44 (6.8) |
| Anaemia, n (%)^3^ | 15/29 (51.7) | 11/24 (45.8) | 16/29 (55.2) |
| Mild anaemia, n (%)^4^ | 13/29 (44.8) | 6/24 (25.0) | 10/29 (34.5) |
| Moderate anaemia, n (%)^5^ | 2/29 (6.9) | 5/24 (20.8) | 6/29 (20.7) |
| Iron deficiency, n (%)^6^ | 1/33 (3.0) | 0/35 (0) | 0/34 (0) |
| Microcytosis, n (%)^7^ | 0/29 (0) | 0/29 (0) | 1/29 (3.5) |
| Vitamin B12 deficiency, n (%)^8^ | 0/34 (0) | 1/33 (3.0) | 2/41 (4.9) |
| Vitamin A deficiency, n (%)^9^ | 15/34 (44.1) | 6/30 (20.0) | 5/30 (16.7) |
| Inflammation, n (%)^10^ | 4/42 (9.5) | 3/38 (7.9) | 2/43 (4.7) |
| **Micronutrient and inflammation biomarkers**^11^ | | | |
| Haemoglobin (n=105), g/dL | 11.0 (10.6, 11.4) | 11.4 (10.8, 11.9) | 11.2 (10.7, 11.6) |
| Ferritin (n=102), µg/L | 175.7 (143.1, 215.8) | 187.5 (147.5, 238.3) | 162.3 (124.6, 211.3) |
| Serum iron (n=106), µmol/L | 14.8 (13.3, 16.5) | 15.1 (13.6, 16.7) | 15.6 (14.5, 16.9) |
| Transferrin (n=122), g/L | 2.0 (1.9, 2.1) | 2.0 (1.8, 2.1) | 2.1 (2.0, 2.2) |
| TSAT (n=106), % | 28.9 (25.7, 32.6) | 30.6 (26.6, 35.2) | 29.2 (26.9, 31.7) |
| MCV (n=82), fL | 84.1 (81.1, 87.3) | 85.0 (81.5, 88.7) | 85.2 (81.8, 88.7) |
| Vitamin B12 (n=108), pg/mL | 478.9 (433.3, 529.3) | 445.4 (381.1, 520.6) | 461.6 (408.0, 522.1) |
| Vitamin A (n=94), µmol/L | 0.7 (0.7, 0.8) | 0.8 (0.7, 0.9) | 0.8 (0.7, 0.9) |
| Folate (n=108), ng/mL | 12.3 (11.1, 13.6) | 11.8 (10.6, 13.0) | 11.8 (10.8, 12.8) |
| Zinc (n=112), µmol/L | 19.8 (17.4, 22.4) | 21.4 (19.1, 23.9) | 20.5 (18.5, 22.7) |
| α-tocopherol (n=94), µmol/L | 24.7 (21.2, 28.7) | 24.8 (22.4, 27.5) | 25.3 (23.5, 27.2) |
| γ-tocopherol (n=94), µmol/L | 0.8 (0.6, 1.0) | 0.9 (0.8, 1.0) | 0.9 (0.8, 1.0) |
| Haptoglobin (n=122), g/L | 0.4 (0.3, 0.5) | 0.4 (0.3, 0.5) | 0.4 (0.3, 0.5) |
| CRP (n=123), mg/L | 0.9 (0.6, 1.3) | 1.0 (0.7, 1.3) | 0.8 (0.6, 1.1) |

**Abbreviations**: TSAT denotes transferrin saturation; MCV, mean corpuscular volume; IQR, interquartile range; and CRP, C-reactive protein. ^1^Only participants with measured micronutrient biomarkers are included in this analysis. Folate deficiency was not observed in this cohort. ^2^Weight-for-age z scores <-2; ^3^Haemoglobin <11.0 g/dL; ^4^Haemoglobin 10.0 – 10.9 g/dL; ^5^Haemoglobin 7.0 – 9.9 g/dL; ^6^Plasma ferritin <12μg/L or <30μg/L in the presence of inflammation; ^7^MCV <67 fL; ^8^Vitamin B12 <200 pg/mL; ^9^Retinol <0.7 µmol/L; ^10^C-reactive protein >5mg/L; ^11^Geometric means and 95% confidence intervals are presented.

**Table S5.** Baseline characteristics of study participants in the PATH-wSP vaccine trial

|  | **Normal saline,** ^1^  n=86 | **0.6 mg PATH-wSP,** ^1^  n=47 | **1 mg PATH-wSP,** ^1^  n=36 |
| --- | --- | --- | --- |
| Median age, months (IQR) | 16.0 (14.0, 18.0) | 15.0 (14.0, 19.0) | 16.5 (14.5, 18.0) |
| Sex, male n (%) | 40/86 (46.5) | 26/47 (55.3) | 14/36 (38.9) |
| Underweight, n (%)^2^ | 17/82 (19.8) | 13/42 (27.7) | 2/36 (5.6) |
| Anaemia, n (%)^3^ | 57/82 (69.5) | 26/41 (63.4) | 18/36 (50.0) |
| Mild anaemia, n (%)^4^ | 34/82 (41.5) | 13/41 (31.7) | 8/36 (22.2) |
| Moderate anaemia, n (%)^5^ | 23/82 (28.1) | 13/41 (31.7) | 10/36 (27.8) |
| Iron deficiency, n (%)^6^ | 43/85 (50.6) | 19/47 (40.4) | 19/47 (52.8) |
| Iron deficiency anaemia, n (%)^7^ | 32/81 (39.5) | 15/41 (36.6) | 11/36 (30.6) |
| Microcytosis, n (%)^8^ | 51/82 (62.2) | 30/41 (73.2) | 20/36 (55.6) |
| Vitamin B12 deficiency, n (%)^9^ | 2/70 (2.9) | 0/39 (0) | 3/31 (9.7) |
| Vitamin A deficiency, n (%)^10^ | 10/74 (13.5) | 3/34 (8.8) | 4/33 (12.1) |
| Inflammation, n (%)^11^ | 24/86 (27.9) | 12/47 (25.5) | 9/36 (25.0) |
| **Micronutrient and inflammatory biomarkers**^12^ | | | |
| Haemoglobin (n=159), g/dL | 10.1 (9.8, 10.3) | 10.1 (9.7, 10.5) | 10.2 (9.9, 10.5) |
| Ferritin (n=168), µg/L | 15.6 (13.3, 18.3) | 14.4 (11.8, 17.4) | 13.9 (10.9, 17.6) |
| Serum iron (n=168), µmol/L | 6.4 (5.9, 6.9) | 6.4 (5.7, 7.3) | 6.5 (5.6, 7.4) |
| Transferrin (n=169), g/L | 3.2 (3.1, 3.3) | 3.2 (3.1, 3.3) | 3.2 (3.1, 3.3) |
| TSAT (n=168), % | 8.0 (7.3, 8.8) | 8.0 (7.1, 9.2) | 8.1 (7.0, 9.3) |
| MCV (n=159), fL | 64.2 (62.9, 65.5) | 62.8 (60.7, 64.9) | 65.7 (63.7, 67.9) |
| Vitamin B12 (n=140), pg/mL | 462.7 (418.6, 511.5) | 426.4 (370.1, 491.2) | 422.5 (359.0, 497.3) |
| Vitamin A (n=141), µmol/L | 0.9 (0.9, 1.0) | 0.9 (0.8, 1.0) | 0.9 (0.8, 1.0) |
| Folate (n=140), ng/mL | 12.6 (11.7, 13.6) | 11.8 (10.8, 12.9) | 14.1 (12.6, 15.6) |
| Zinc (n=163), µmol/L | 16.7 (16.2, 17.3) | 17.6 (16.4, 18.8) | 16.2 (15.3, 17.1) |
| α-tocopherol (n=141), µmol/L | 21.8 (20.7, 22.9) | 23.8 (21.8, 25.9) | 19.8 (18.2, 21.7) |
| γ-tocopherol (n=141), µmol/L | 0.8 (0.7, 0.9) | 0.8 (0.6, 0.9) | 0.6 (0.5, 0.7) |
| Haptoglobin (n=169), g/L | 1.0 (0.9, 1.1) | 1.1 (1.0, 1.2) | 1.1 (0.9, 1.2) |
| CRP (n=169), mg/L | 3.0 (2.3, 3.8) | 2.6 (1.9, 3.6) | 2.8 (2.2, 3.7) |

**Abbreviations**: TSAT denotes transferrin saturation; MCV, mean corpuscular volume; IQR, interquartile range; CRP, and C-reactive protein.^1^Only participants with micronutrient biomarker status included. Folate deficiency was not observed in this cohort; ^2^Weight-for-age z scores <-2; ^3^Haemoglobin <11.0 g/dL; ^4^Haemoglobin 10.0 – 10.9 g/dL; ^5^Haemoglobin 7.0 – 9.9 g/dL; ^6^Plasma ferritin <12μg/L or <30μg/L in the presence of inflammation; ^7^Iron deficiency and anaemia; ^8^MCV <67 fL; ^9^Vitamin B12 <200 pg/mL; ^10^Retinol <0.7 µmol/L; ^11^C-reactive protein >5mg/L; ^12^Geometric means and 95% confidence intervals are presented.

**Table S6.** Associations between baseline micronutrient status and composite IgG z-scores in the primary full-dose groups

|  | **PCV10 (PRISM)** ^1^ | | | **Full–dose PCV10 (FPCV)** ^2^ | | | **Full dose PCV13 (FPCV)** ^2^ | | | **1 mg PATH–wSP** ^2^ | | |
| --- | --- | --- | --- | --- | --- | --- | --- | --- | --- | --- | --- | --- |
|  | **n/N (%)** | **Adj. β (95% CI)** ^3^ | **P** ^3^ | **n/N (%)** | **Adj. β (95% CI)** ^3^ | **P** ^3^ | **n/N (%)** | **Adj. β (95% CI)** ^3^ | **P** ^3^ | **n/N (%)** | **Adj. β (95% CI)** ^3^ | **P** ^3^ |
| Anaemia | NA^4^ | NA^4^ | NA^4^ | 12/32 (37.5) | 0.4 (–0.3, 1.1) | 0.16 | 15/29 (51.7) | –0.4 (–1.0, 0.2) | 0.26 | 18/36 (50.0) | –0.8 (–1.2, –0.4) | 0.0001 |
| Mild anaemia | NA^4^ | NA^4^ | NA^4^ | 7/32 (21.9) | 0.4 (–0.4, 1.2) | 0.24 | 13/29 (44.8) | –0.4 (–1.0, 0.3) | 0.30 | 8/36 (22.2) | –0.8 (–1.3, –0.3) | 0.001 |
| Moderate anaemia | NA^4^ | NA^4^ | NA^4^ | 5/32 (15.6) | 0.3 (–0.7, 1.3) | 0.57 | 2/29 (6.9) | –0.5 (–1.9, 0.9) | 0.61 | 10/36 (27.8) | –0.8 (–1.2, –0.3) | 0.002 |
| Vitamin B12 deficiency | 4/101 (4.0) | –0.9 (–1.4, –0.3) | 0.002 | 0/41 | NA^5^ | NA^5^ | 0/34 | NA^5^ | NA^5^ | 3/31 (9.7) | –0.2 (–1.1, 0.7) | 0.69 |
| Iron deficiency | 21/91 (23.1) | 0.1 (–0.2, 0.4) | 0.59 | 0/42 | NA^5^ | NA^5^ | 1/33 (3.0) | NA^5^ | NA^5^ | 11/36 (30.6) | –0.3 (–0.8, 0.2) | 0.18 |
| Iron deficiency anaemia | NA^4^ | NA^4^ | NA^4^ | 0/32 | NA^5^ | NA^5^ | 0/29 | NA^5^ | NA^5^ | 11/36 (30.6) | –0.8 (–1.3, –0.2) | 0.008 |
| Vitamin A deficiency | 33/95 (34.7) | –0.2 (–0.5, 0.1) | 0.17 | 10/35 (28.6) | 0.5 (–0.2, 1.1) | 0.13 | 15/34 (44.1) | 0.2 (–0.3, 0.7) | 0.45 | 4/33 (12.1) | 0.1 (–0.7, 0.9) | 0.78 |
| Folate deficiency | 16/101 (15.8) | 0.2 (–0.1, 0.5) | 0.15 | 0/41 | NA^5^ | NA^5^ | 0/34 | NA^5^ | NA^5^ | 0/31 | NA^5^ | NA^5^ |
| Vitamin D deficiency | 1/101 (1.6) | NA^5^ | NA^5^ | NA^4^ | NA^4^ | NA^4^ | NA^4^ | NA^4^ | NA^4^ | NA^4^ | NA^4^ | NA^4^ |

^1^In the PRISM trial, Groups A and B were combined because both received their first PCV10 dose at baseline. ^2^Only primary groups receiving full vaccine doses are shown. ^3^Adjusted β coefficients and P values were estimated using multivariable linear regression models of composite IgG z scores and log-transformed biomarkers (except haemoglobin) adjusted for age, sex, underweight status, and log-transformed C-reactive protein concentrations. ^4^Biomarker not measured in the corresponding trial. ^5^Prevalence was too low for regression analysis. NA; not available.

**Table S7.** Associations between baseline micronutrient status and composite IgG z-scores in the fractional dose FPCV and PATH-wSP groups

|  | **40% PCV10 (FPCV)** | | **20% PCV10 (FPCV)** | | **40% PCV13 (FPCV)** | | **20% PCV13 (FPCV)** | | **0.6 mg PATH–wSP** | |
| --- | --- | --- | --- | --- | --- | --- | --- | --- | --- | --- |
|  | **Adj. β (95% CI)** **^1^** | **P ^1^** | **Adj. β (95% CI)** **^1^** | **P ^1^** | **Adj. β (95% CI)** **^1^** | **P ^1^** | **Adj. β (95% CI)** **^1^** | **P ^1^** | **Adj. β (95% CI)** **^1^** | **P ^1^** |
| Haemoglobin, g/dL | 0.1 (–0.1, 0.3) | 0.41 | –0.1 (–0.3, 0.0) | 0.14 | –0.1 (–0.3, 0.0) | 0.11 | –0.1 (–0.3, 0.0) | 0.12 | 0.0 (–0.3, 0.2) | 0.70 |
| Ferritin, µg/L | 0.2 (–0.4, 0.7) | 0.53 | –0.1 (–0.5, 0.2) | 0.49 | 0.0 (–0.3, 0.4) | 0.78 | 0.2 (–0.1, 0.5) | 0.18 | –0.1 (–0.5, 0.3) | 0.61 |
| Serum iron, µmol/L | 0.6 (–0.4, 1.6) | 0.25 | –0.2 (–1.1, 0.6) | 0.56 | 0.1 (–0.8, 1.0) | 0.78 | –0.0 (–1.2, 1.2) | 0.95 | 0.0 (–0.8, 0.7) | 0.91 |
| Transferrin, g/L | –0.3 (–0.9, 0.4) | 0.41 | 0.2 (–0.2, 0.7) | 0.29 | 0.2 (–0.3, 0.8) | 0.35 | –0.1 (–0.7, 0.5) | 0.73 | –0.1 (–0.7, 0.5) | 0.71 |
| TSAT, % | 0.5 (–0.3, 1.3) | 0.23 | –0.2 (–0.8, 0.4) | 0.45 | –0.1 (–0.7, 0.4) | 0.63 | 0.2 (–0.9, 1.2) | 0.74 | 0.0 (–0.6, 0.7) | 0.97 |
| Vitamin B12, pg/mL | –0.5 (–1.2, 0.2) | 0.20 | –0.8 (–1.5, –0.1) | 0.03 | 0.3 (–0.2, 0.8) | 0.18 | –0.3 (–0.9, 0.3) | 0.27 | –0.1 (–0.8, 0.6) | 0.79 |
| Vitamin A, µmol/L | –0.2 (–2.0, 1.7) | 0.84 | –0.2 (–1.1, 0.7) | 0.64 | 0.7 (–0.3, 1.8) | 0.17 | –0.0 (–1.0, 0.9) | 0.95 | 0.0 (–1.6, 1.6) | 0.98 |
| Folate, ng/mL | –0.1 (–1.0, 0.9) | 0.86 | 0.5 (–0.3, 1.3) | 0.22 | 0.4 (–0.4, 1.1) | 0.34 | –0.8 (–1.6, –0.1) | 0.03 | 0.3 (–0.7, 1.3) | 0.55 |
| Zinc, µmol/L | –0.1 (–0.9, 0.7) | 0.83 | –0.1 (–0.6, 0.4) | 0.75 | 0.3 (–0.5, 1.0) | 0.47 | –0.2 (–1.0, 0.5) | 0.51 | –0.1 (–1.4, 1.1) | 0.84 |
| Vitamin E, µmol/L | –0.4 (–2.7, 1.9) | 0.71 | –0.2 (–0.9, 0.4) | 0.51 | 0.3 (–0.8, 1.3) | 0.60 | –1.3 (–2.5, –0.1) | 0.04 | 0.2 (–1.0, 1.5) | 0.72 |

^1^Adjusted regression coefficients (β) and P values were estimated using multivariable linear regression models of composite IgG z scores and log-transformed biomarkers (except haemoglobin and transferrin) adjusted for age, sex, underweight status, and log-transformed C-reactive protein concentrations.

**Table S8.** Summary of associations between baseline micronutrient biomarker levels and vaccine antibody responses

| **Vaccine trial** | **Vaccine group** | **Micronutrient** | **Associated antigen(s)/ serotype(s)** | **Direction of association^1^** |
| --- | --- | --- | --- | --- |
| PATH-wSP | 1 mg PATH-wSP | Haemoglobin^2^ | L460D, BCH0785, StkP, PiuA | Increased IgG |
| PRISM | Group A & B | Vitamin B12^2^ | STs 1, 6B | Increased IgG |
|  |  |  | STs 1, 4, 14 | Increased OPA |
| FPCV | Full PCV10 dose | Vitamin B12 | ST 9V | Increased IgG |
|  | 40% PCV10 dose | Ferritin | ST 7F | Increased IgG |
|  | 20% PCV10 dose | Haemoglobin | ST 18C | Lower IgG |
|  |  | Vitamin B12 | ST 6B | Lower IgG |
|  | Full PCV13 dose | Ferritin | STs 1, 6A, 9V | Increased IgG |
|  |  | Folate | ST 5 | Lower IgG |
|  |  | Vitamin B12 | ST 6B | Increased IgG |
|  | 40% PCV13 dose | Haemoglobin | ST 14 | Increased IgG |
|  |  | Vitamin A | ST 6A | Increased IgG |
|  | 20% PCV13 dose | Folate | STs 4, 5, 7F, 9V, 19F | Lower IgG |
|  |  | Vitamin B12 | ST 6B | Lower IgG |
|  |  | Vitamin E | ST 4 | Lower IgG |

^1^Based on linear regression models adjusting for age, sex, underweight, and log-transformed CRP levels. ^2^Statistically significant after Benjamini–Hochberg correction of multiple testing.

**Supplementary Figures**

**Figure S1.** Serotype-specific post-vaccination IgG responses across the three vaccine trials

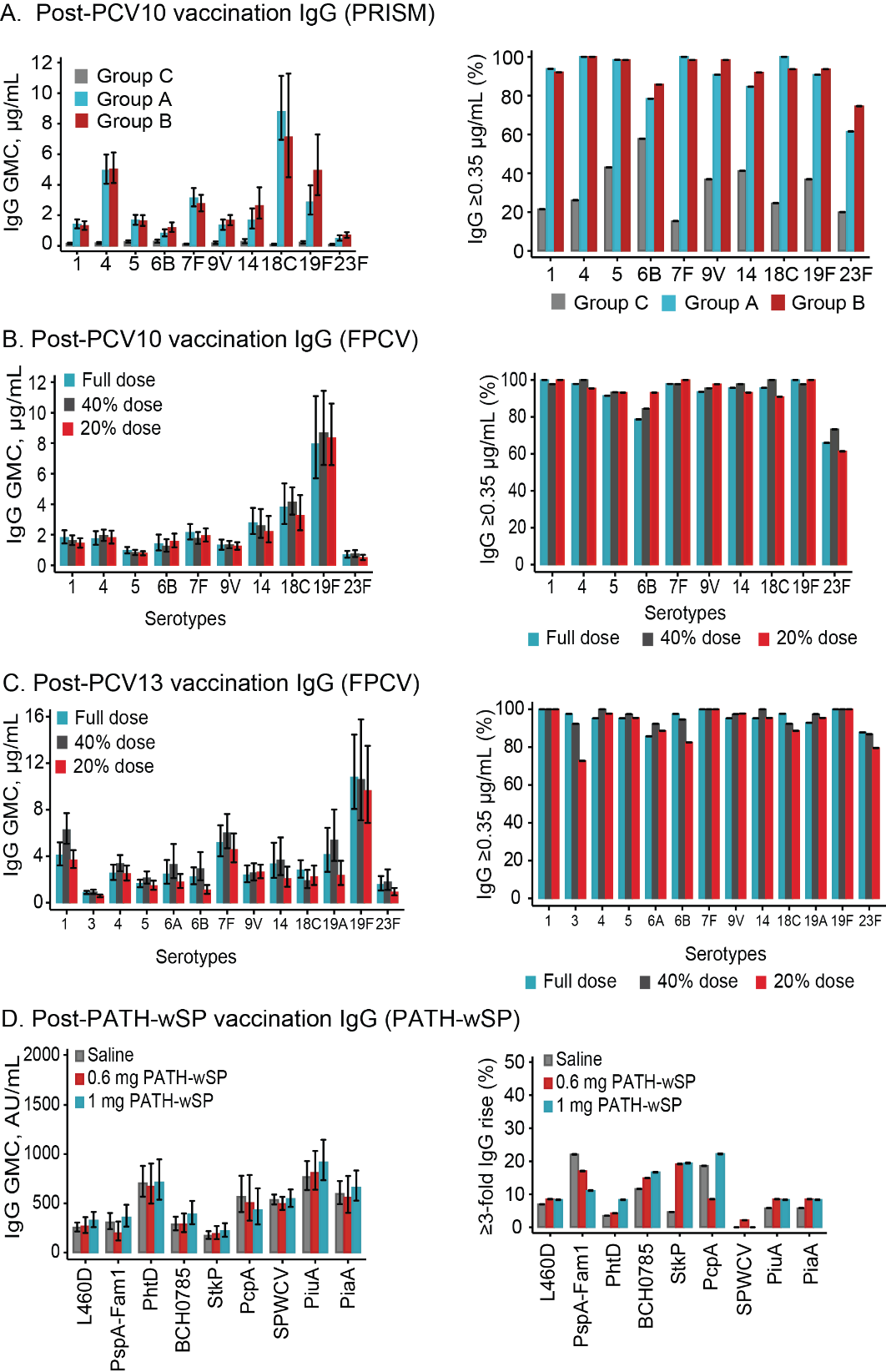

Serotype- or antigen-specific IgG geometric mean concentrations (GMCs) and proportion of participants achieving IgG concentrations ≥0.35 µg/mL or with ≥fold IgG rise following vaccination with A) one dose of PCV10 or placebo (PRISM: Group A, blue bars; Group B, red bars; Group C, grey bars); B) two primary doses of PCV13 or C) PCV10 (FPCV: full dose, blue bars; 40% dose, grey bars; 20% dose, red bars); or D) two doses of PATH-wSP or control (1 mg, blue bars; 0.6 mg, red bars; saline, grey bars). Only participants with available baseline micronutrient biomarker data were included in the analyses.

**Figure S2**. Serotype-specific opsonophagocytic activity in the PRISM vaccine trial

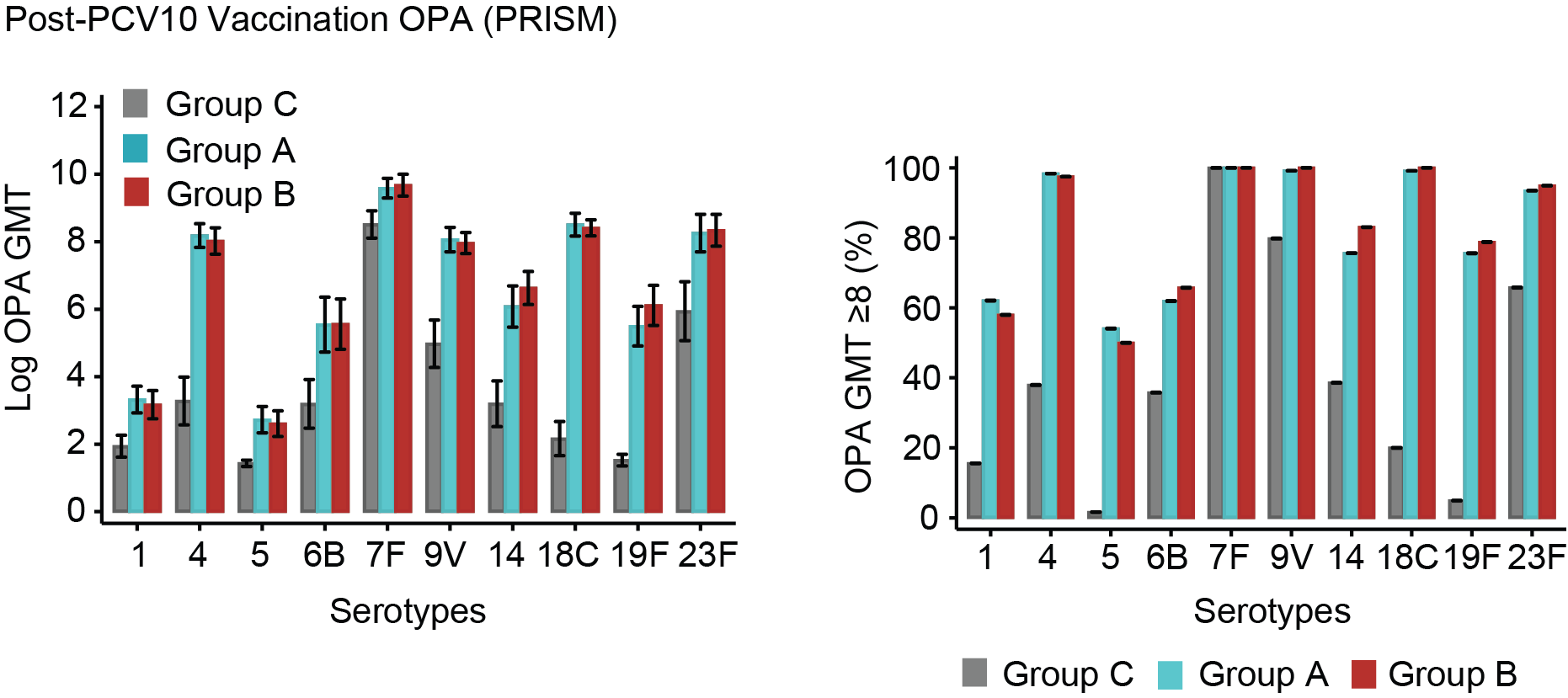

A) Opsonophagocytic activity (OPA) geometric mean titres (GMTs) and B) proportion of participants with an OPA GMT ≥8 in participants receiving PCV10 vaccine (Group A, blue bars; and Group B, red bars) or placebo vaccines (Group C, grey bars). OPA GMTs were assessed in sera collected 30 days post one vaccine dose. Error bars represent 95% confidence intervals. Only participants with available baseline micronutrient biomarker data were included in the analysis.

**Figure S3**. Association between baseline biomarkers of micronutrient status and IgG responses to pneumococcal antigens following 1 mg PATH-wSP vaccination

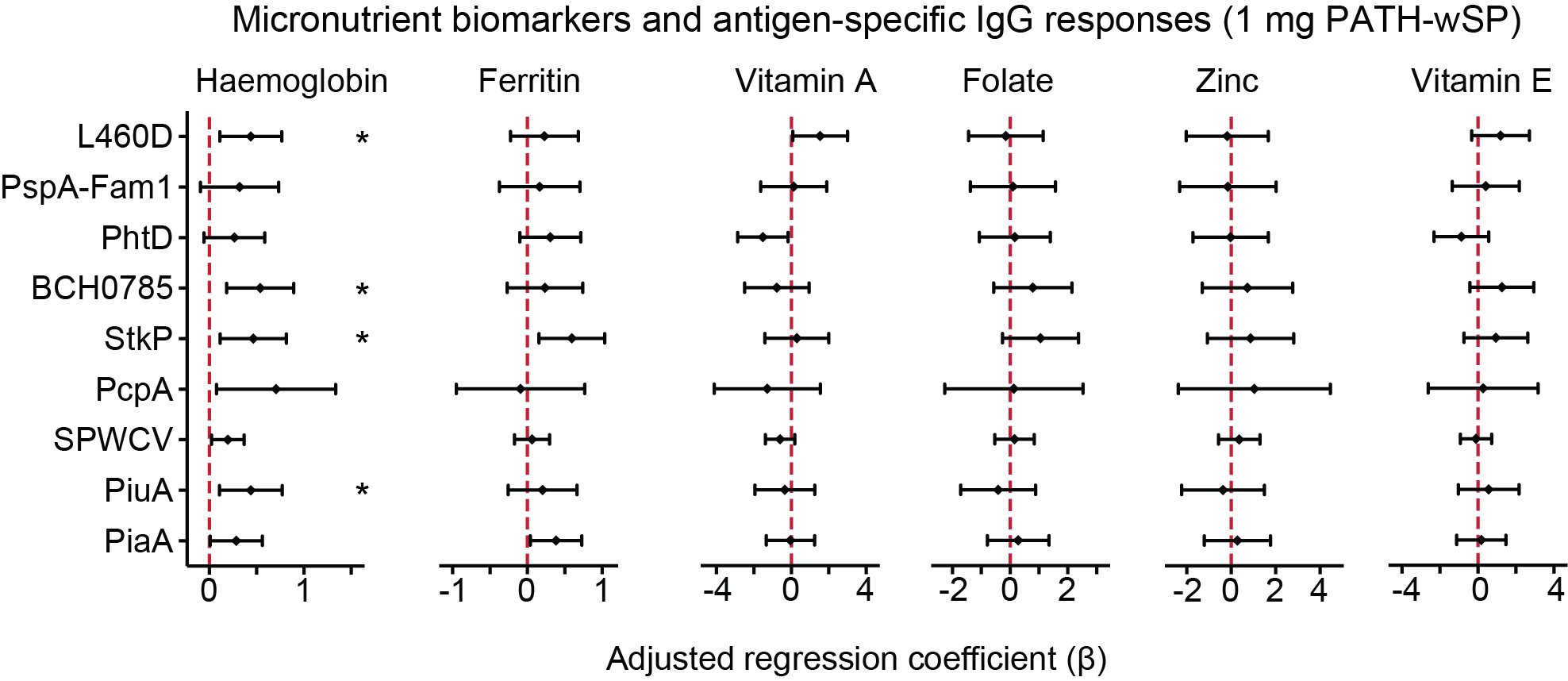

IgG geometric mean concentrations were assessed 28 days after the second vaccine dose. Adjusted regression coefficients (β) were derived from linear regression models adjusting for age, sex, underweight status, and log-transformed C-reactive protein levels. Vitamin E status was assessed using α-tocopherol concentrations. Statistical significance after Benjamini-Hochberg correction for multiple antibody comparisons: * denotes P<0.05; **, P<0.01; and ***, P<0.001.

**Figure S4.** Post-vaccination IgG concentrations among participants receiving 1 mg PATH-wSP vaccine, stratified by anaemia status

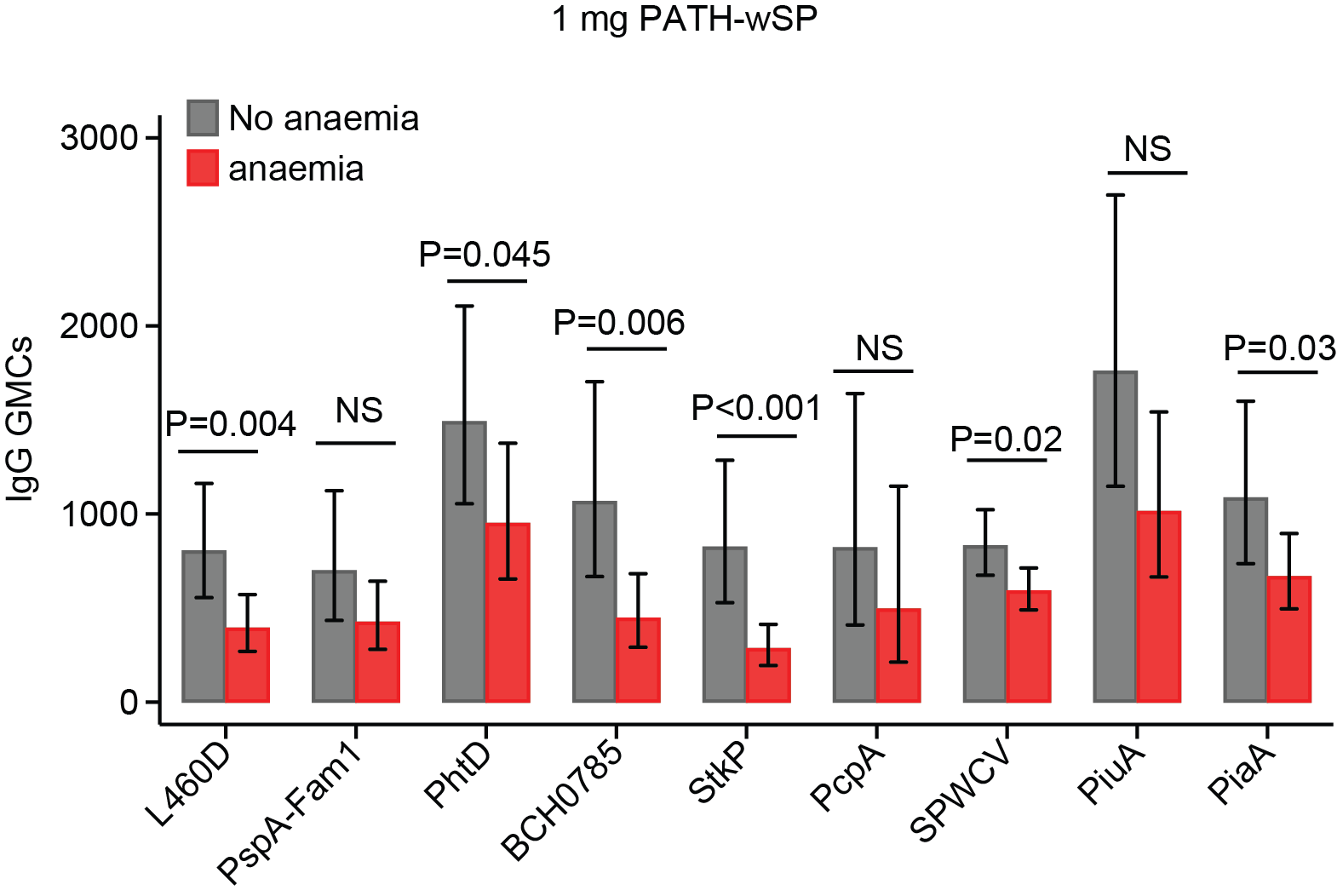

P values were derived from linear regression models adjusted for age, sex, underweight status, and log-transformed C-reactive protein levels. NS denotes not significant (P≥0.05).

**Figure S5**. Associations between baseline anaemia and serotype- or antigen-specific IgG responses.

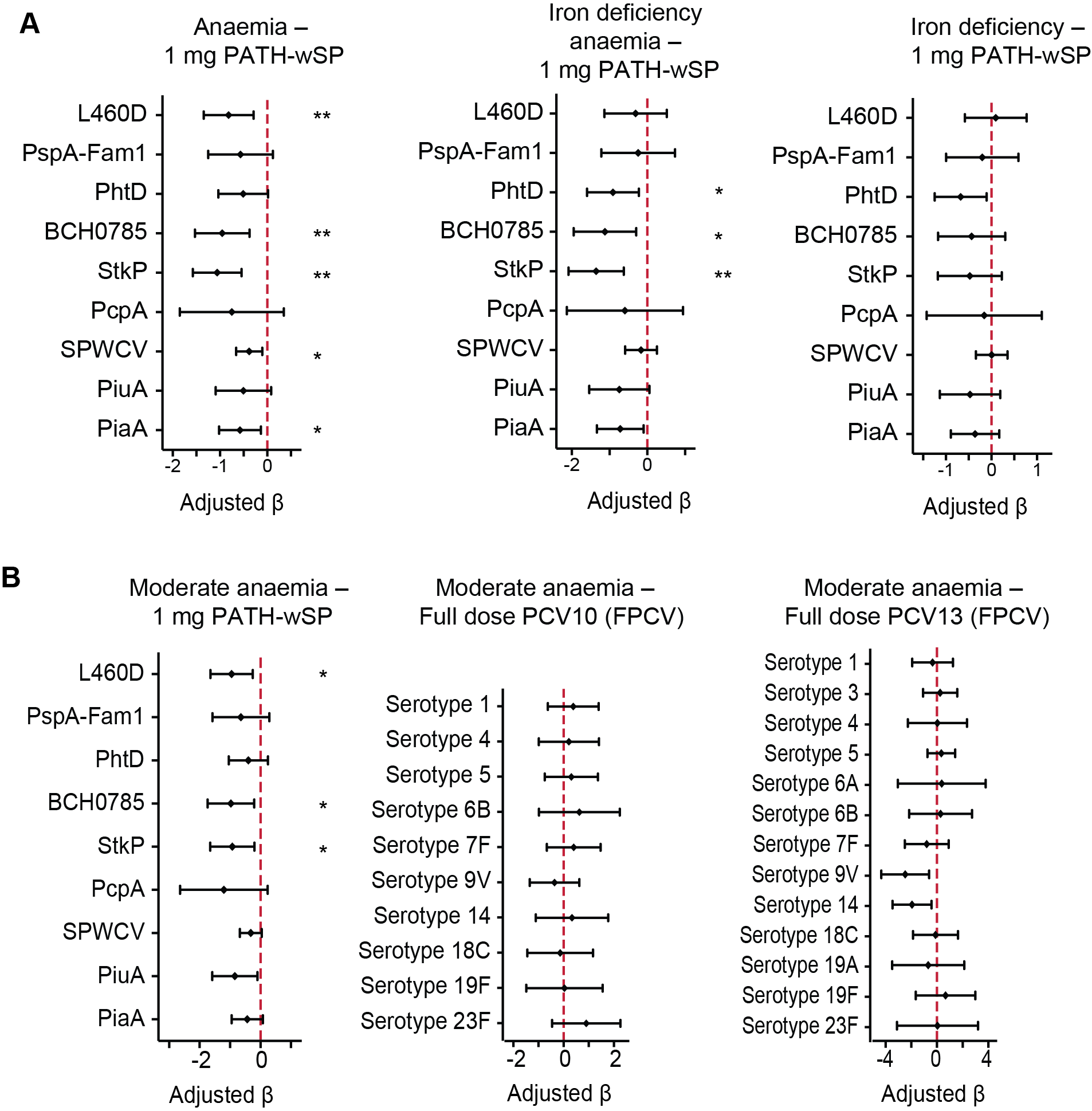

A) Associations with anaemia, iron deficiency anaemia, and iron deficiency among recipients of 1 mg PATH-wSP vaccine. B) Associations with moderate anaemia following two doses of 1 mg PATH-wSP vaccine or two primary full doses of PCV10 or PCV13 (FPCV trial). IgG responses were measured in serum samples collected 28 days after vaccination. Adjusted β coefficients were estimated using linear regression models adjusted for age, sex, underweight status, and log-transformed C-reactive protein concentrations. Statistical significance after Benjamini-Hochberg correction for multiple testing: * denotes P<0.05; **, P<0.01; and ***, P<0.001. Haemoglobin concentrations were not available in the PRISM trial.

**Figure S6**. Association between baseline micronutrient biomarker levels and pneumococcal IgG responses in recipients of full dose PCV13 and PCV10 in the FPCV vaccine trial

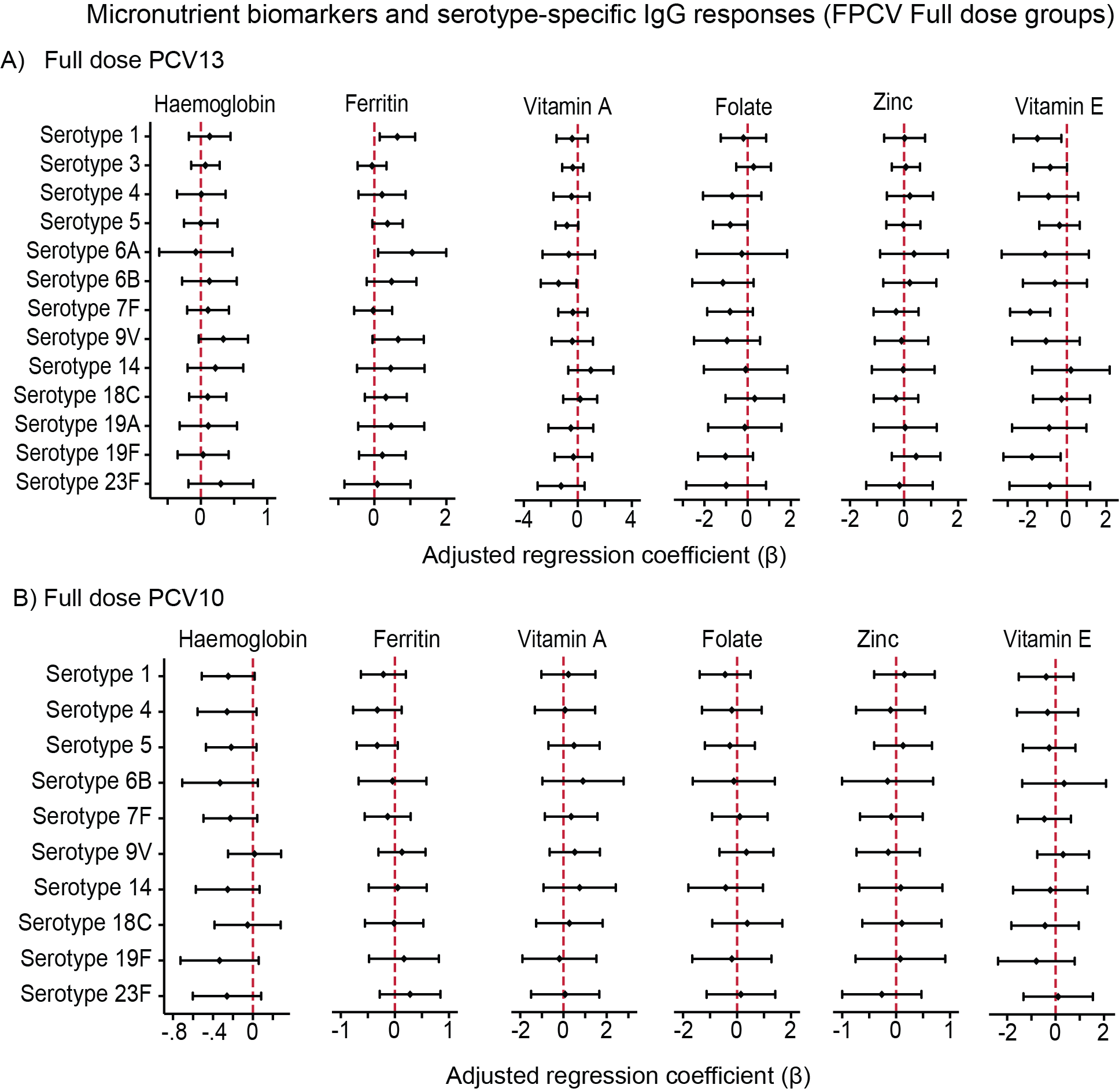

IgG responses were measured 28 days after two primary full doses of A) PCV13 and B) PCV10 vaccination. Adjusted regression coefficients (β) were derived from linear regression models adjusting for age, sex, underweight status, and log-transformed C-reactive protein levels. Statistical significance after Benjamini-Hochberg correction for multiple antibody comparisons: * denotes P<0.05; **, P<0.01; and ***, P<0.001.

**Figure S7**. Association between baseline biomarkers of micronutrient status and pneumococcal IgG responses in the FPCV trial following primary fractional doses of PCV13 and PCV10

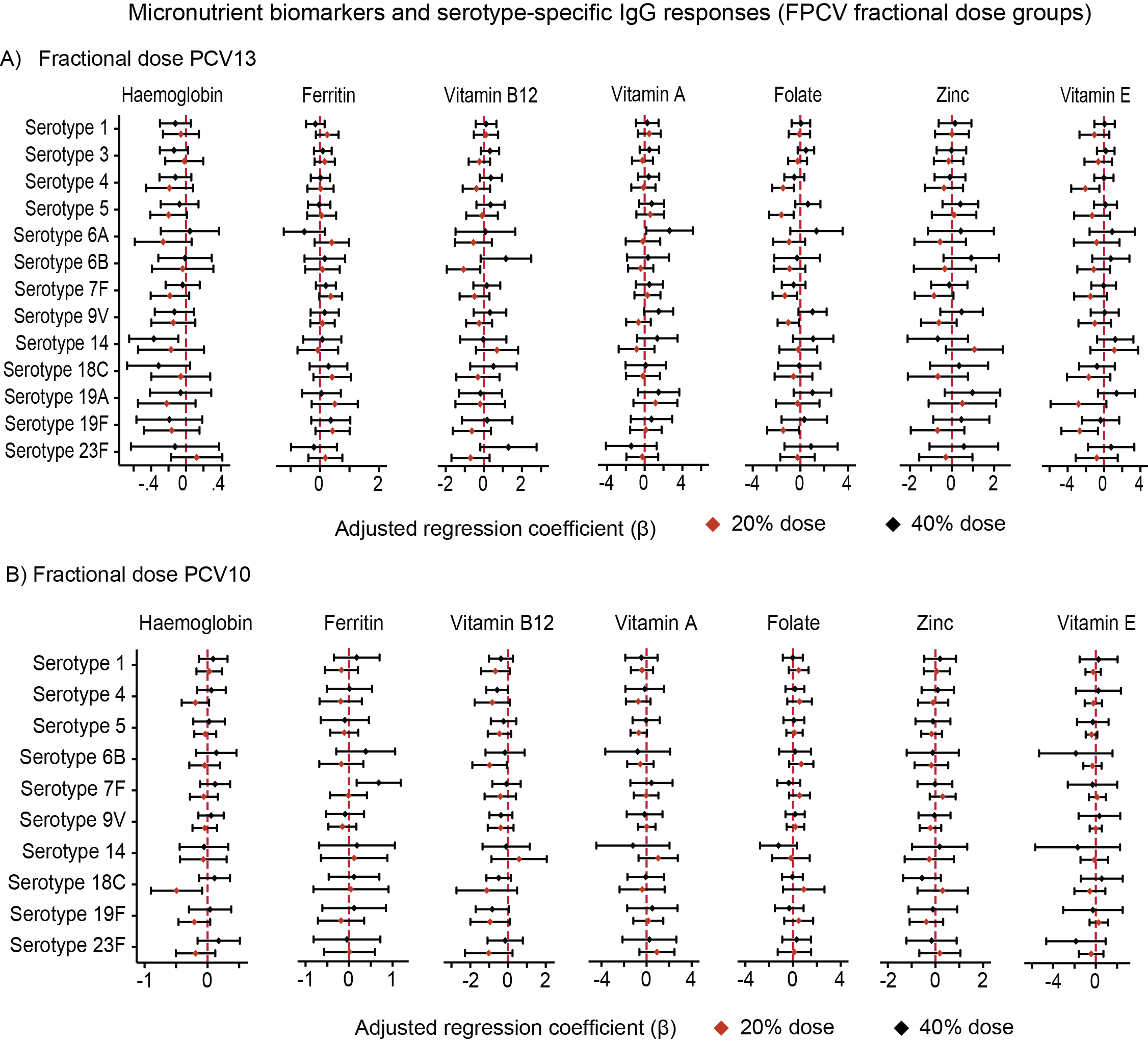

IgG responses were measured 28 days after two primary full doses of A) PCV13 and B) PCV10 vaccination. Black diamond denotes 40% dose; and blue diamond, 20% dose. Adjusted beta coefficients were derived from linear regression models adjusting for age, sex, underweight status, and log-transformed C-reactive protein levels. Statistical significance after Benjamini-Hochberg correction for multiple antibody comparisons: * denotes P<0.05; **, P<0.01; and ***, P<0.001.

**Figure S8.** Associations between baseline vitamin B12 concentrations and pneumococcal opsonophagocytic activity responses following a single PCV10 dose in the PRISM trial

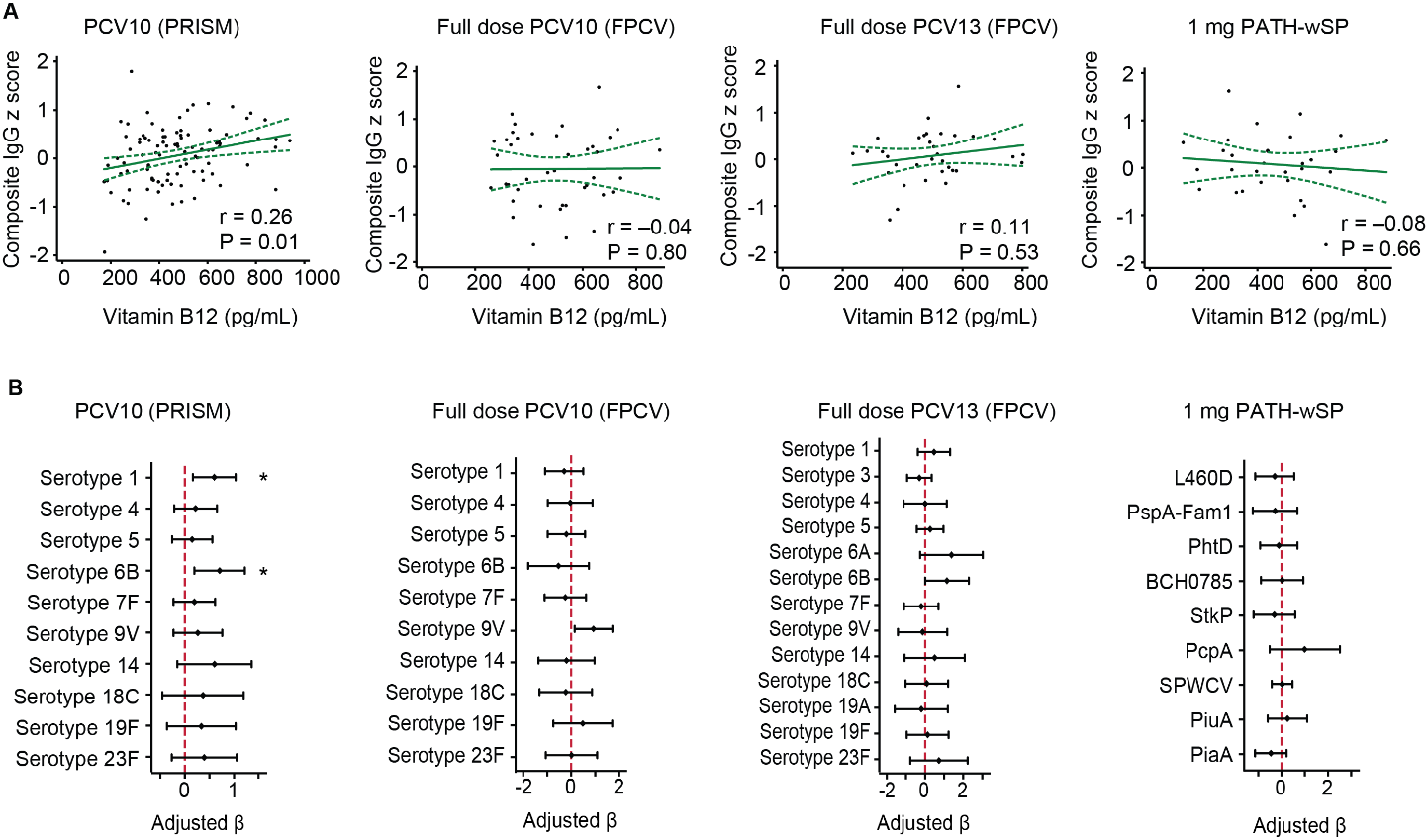

Associations between pneumococcal vaccine antibody responses and baseline vitamin B12 concentrations. A) Spearman rank correlations between baseline haemoglobin concentrations and post-vaccination composite IgG z-scores among participants receiving one dose of PCV10 (PRISM), full doses of PCV10 or PCV13 in the FPCV trial, or 1 mg PATH-wSP. B) Associations between post-vaccination serotype- or antigen-specific IgG responses and baseline vitamin B12 concentrations among participants receiving one dose of PCV10 (PRISM), full doses of PCV10 or PCV13 in the FPCV trial, or 1 mg PATH-wSP. Adjusted β coefficients were estimated using linear regression models adjusted for age, sex, underweight status, and log-transformed C-reactive protein concentrations. Statistical significance after Benjamini-Hochberg correction for multiple antibody comparisons: * denotes P<0.05; **, P<0.01; and ***, P<0.001.

**Figure S9**. Association between baseline micronutrient status and pneumococcal vaccine responses following a single PCV10 dose in the PRISM trial

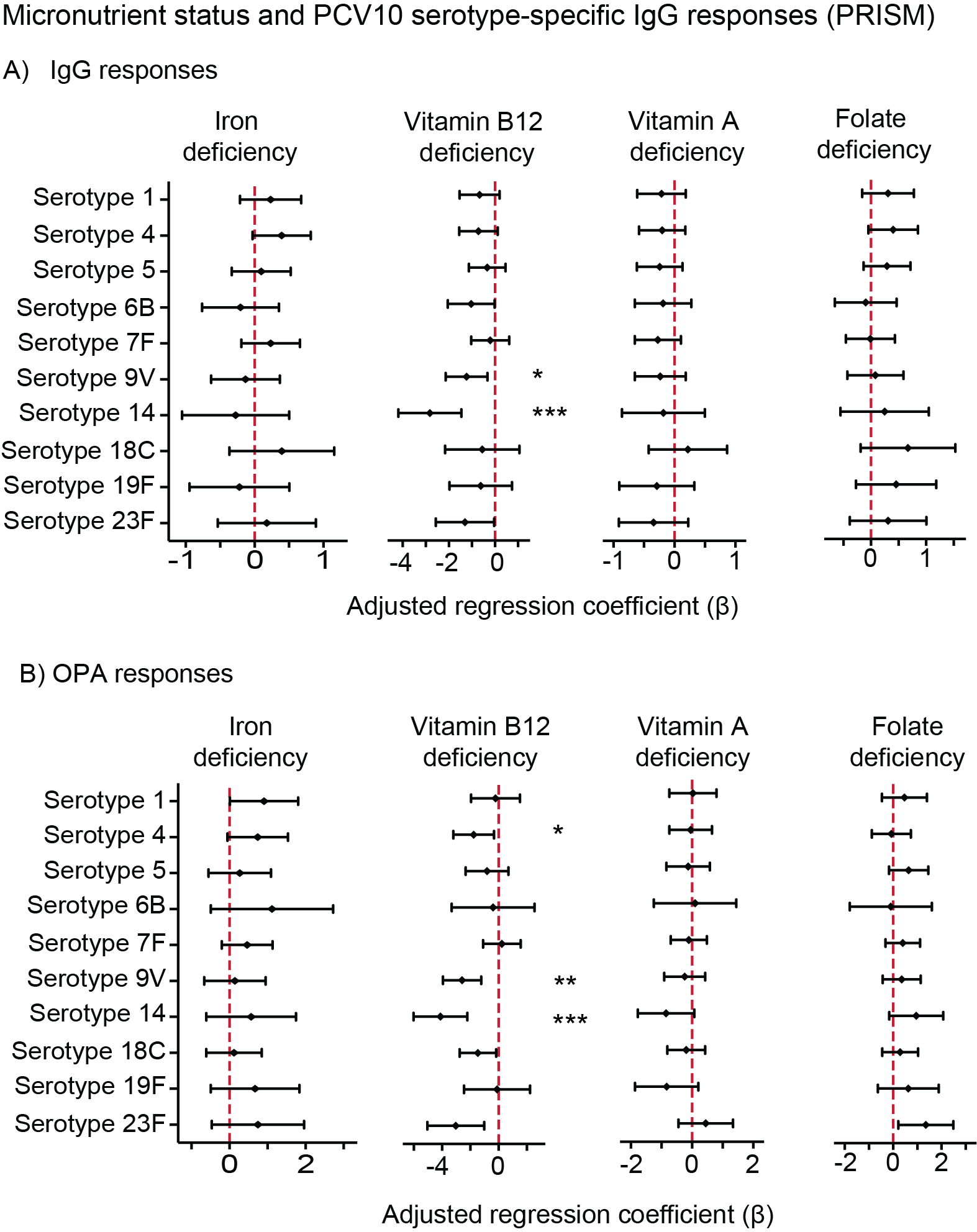

Association between baseline micronutrient status and A) IgG responses or B) opsonophagocytic activity (OPA) responses after one PCV10 dose. Iron deficiency was defined as plasma ferritin <12 μg/L or <30 μg/L in the presence of inflammation; folate deficiency as serum folate <4 ng/mL; vitamin B12 deficiency as serum vitamin B12 <200 pg/mL; and vitamin A deficiency as serum retinol concentrations <0.7 μmol/L. Blue diamond, Group A; and green diamond, Group B. Adjusted beta coefficients were derived from linear regression models adjusting for age, sex, underweight status, and log-transformed C-reactive protein levels. Statistical significance after Benjamini-Hochberg correction for multiple antibody comparisons: * denotes P<0.05; **, P<0.01; and ***, P<0.001.

**Figure S10.** Associations between baseline micronutrient biomarkers and pneumococcal vaccine responses following a single PCV10 dose in the PRISM trial

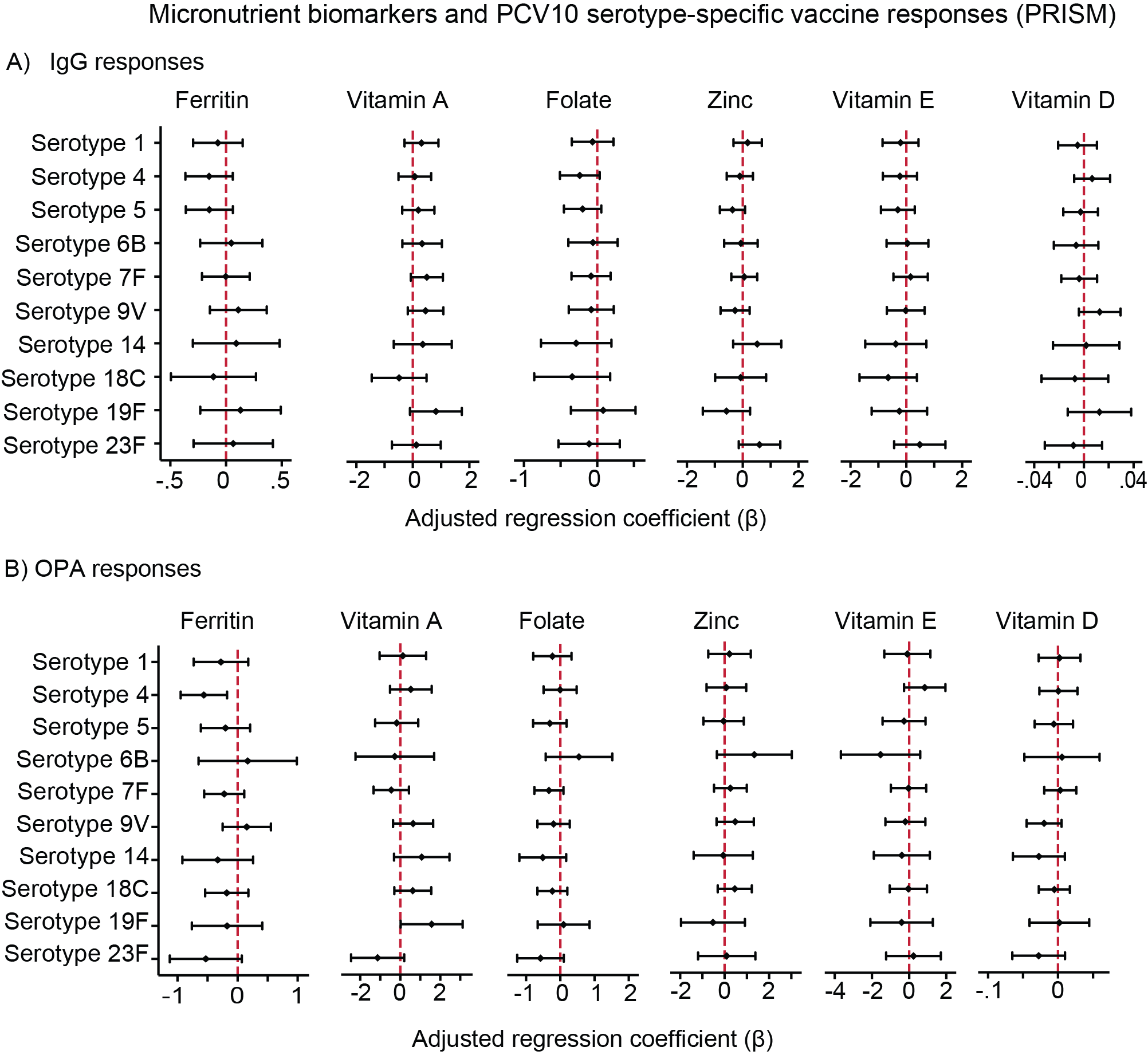

A) Serotype-specific IgG responses and B) Opsonophagocytic activity (OPA) against the ten PCV10 serotypes. Participants in Groups A and B received one dose at baseline and were combined in this analysis. IgG and OPA responses measured 30 days post vaccination. Haemoglobin concentrations were not available in the PRISM trial. Adjusted regression coefficients (β) were derived from linear regression models adjusting for age, sex, underweight status, and log-transformed C-reactive protein levels. Statistical significance after Benjamini-Hochberg correction for multiple antibody comparisons: * denotes P<0.05; **, P<0.01; and ***, P<0.001.

**Figure S11**. Association between baseline micronutrient biomarkers and pneumococcal IgG responses in the FPCV trial among participants receiving three primary PCV10 doses according to the Kenyan Ministry of Health schedule

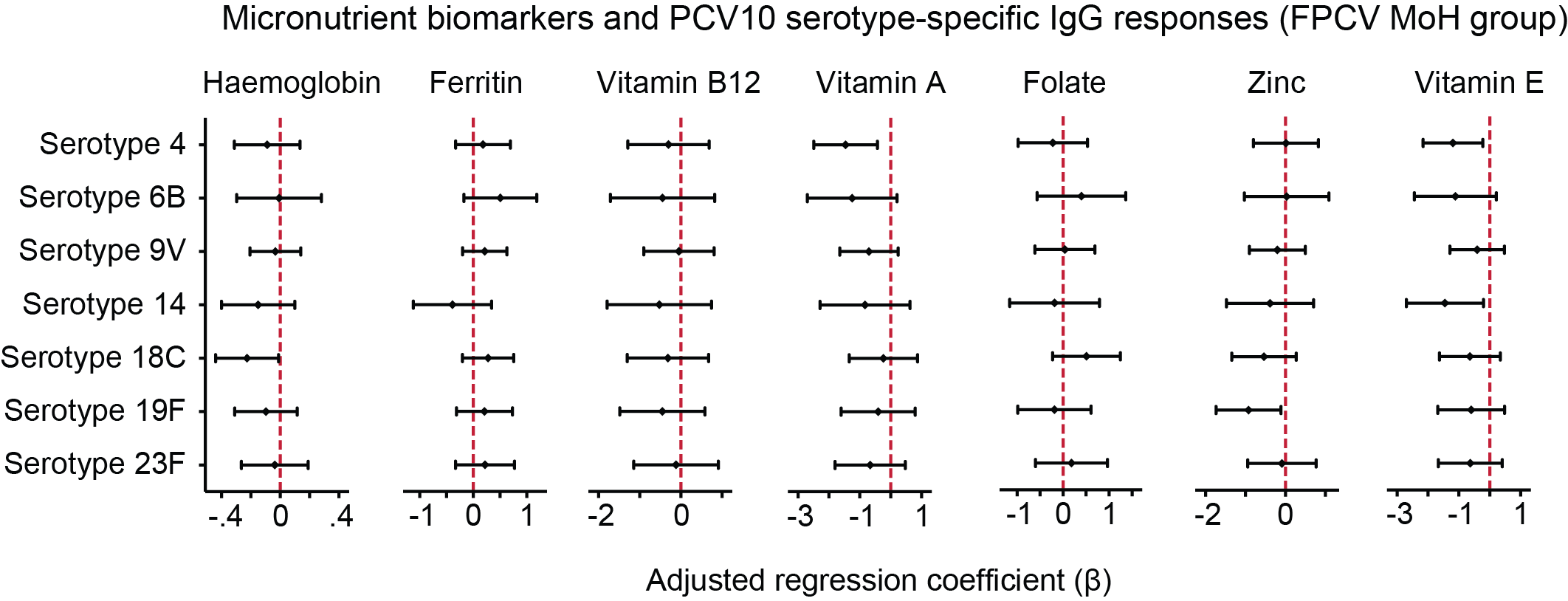

IgG geometric mean concentrations were assessed 28 days post-vaccination. Black diamond denotes 40% dose; and blue diamond, 20% dose. Adjusted beta coefficients were derived from linear regression models adjusting for age, sex, underweight status, and log-transformed C-reactive protein levels. Statistical significance after Benjamini-Hochberg correction for multiple antibody comparisons: * denotes P<0.05; **, P<0.01; and ***, P<0.001.

**Figure S12.** Associations between baseline micronutrient biomarkers and composite IgG z-scores following primary full or fractional PCV10 doses in the FPCV trial

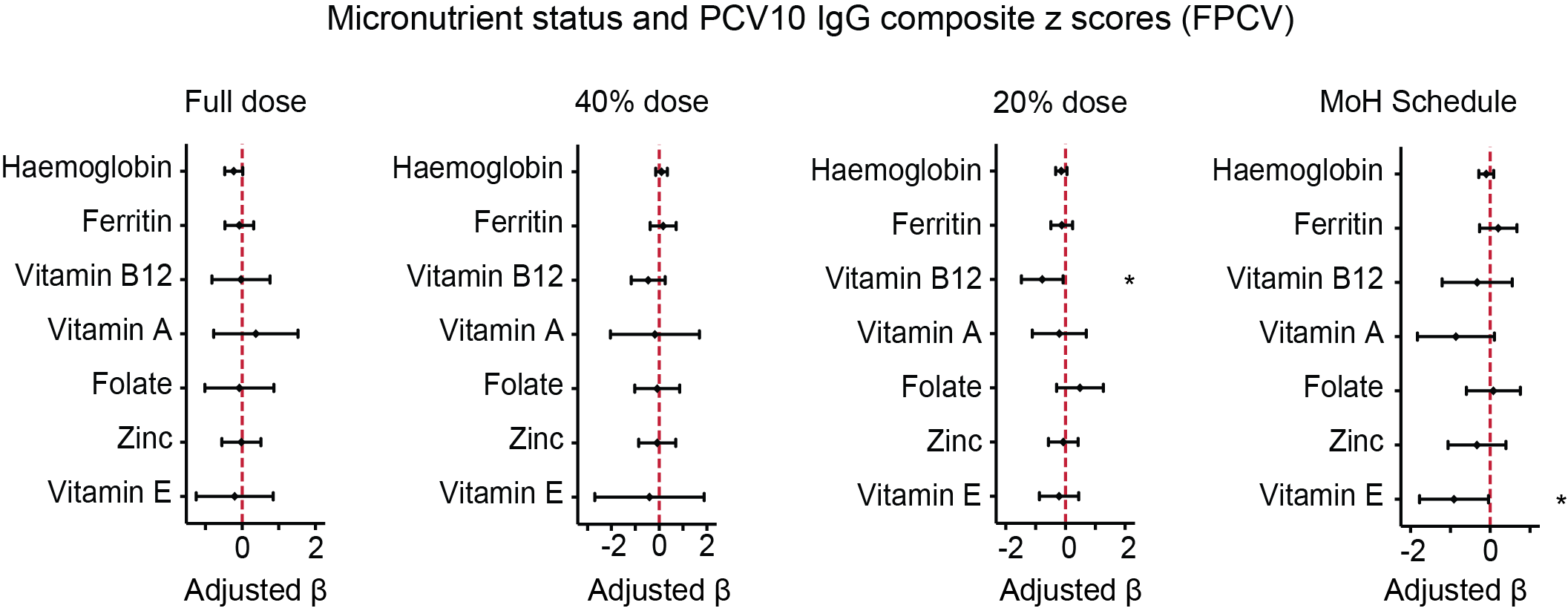

Adjusted β coefficients were estimated using multivariable linear regression models adjusted for age, sex, underweight status, and log-transformed C-reactive protein concentrations. In the group receiving PCV10 according to the Kenyan Ministry of Health (MoH) Expanded Programme on Immunization schedule, composite IgG z-scores were calculated using seven of the ten PCV10 serotypes. Statistical significance: * denotes P<0.05; **, P<0.01; and ***, P<0.001.

**Strobe checklist**

|  | **Item No** | **Recommendation** |
| --- | --- | --- |
| **Title and abstract** | 1 | (*a*) Indicate the study’s design with a commonly used term in the title or the abstract  **This information is provided in the Title and Abstract pages** |
|  |  | (*b*) Provide in the abstract an informative and balanced summary of what was done and what was found  **This information is provided in the Abstract page** |
| **Introduction** | | |
| Background/rationale | 2 | Explain the scientific background and rationale for the investigation being reported  **This information is provided in the Introduction, paragraphs 1 – 2.** |
| Objectives | 3 | State specific objectives, including any prespecified hypotheses  **This information is provided in the Introduction, paragraph 2.** |
| **Methods** | | |
| Study design | 4 | Present key elements of study design early in the paper  **This information is provided in the Methods, subsection on *Study design and participants.*** |
| Setting | 5 | Describe the setting, locations, and relevant dates, including periods of recruitment, exposure, follow-up, and data collection  **This information is provided in Table 1 and in the Methods, subsection on *Study design and participants.*** |
| Participants | 6 | (*a*) Give the eligibility criteria, and the sources and methods of selection of participants  **This information is provided in the Methods, subsection on *Study design and participants.*** |
| Variables | 7 | Clearly define all outcomes, exposures, predictors, potential confounders, and effect modifiers. Give diagnostic criteria, if applicable  **This information is provided in the Methods, subsections on *Procedures and Statistical analyses.*** |
| Data sources/ measurement | 8* | For each variable of interest, give sources of data and details of methods of assessment (measurement). Describe comparability of assessment methods if there is more than one group  **This information is provided in the Methods, subsection on *Procedures.* See also subsection on *Definitions and Supplementary Table S1.*** |
| Bias | 9 | Describe any efforts to address potential sources of bias  **This information is provided in the Methods, subsections on *Statistical analyses.*** |
| Study size | 10 | Explain how the study size was arrived at  **This information is provided in the Methods, subsection on *Study design and participants.* See also subsection on *Statistical analyses.*** |
| Quantitative variables | 11 | Explain how quantitative variables were handled in the analyses. If applicable, describe which groupings were chosen and why |
| Statistical methods | 12 | (*a*) Describe all statistical methods, including those used to control for confounding |
|  |  | (*b*) Describe any methods used to examine subgroups and interactions |
|  |  | (*c*) Explain how missing data were addressed |
|  |  | (*d*) If applicable, describe analytical methods taking account of sampling strategy |
|  |  | (*e*) Describe any sensitivity analyses  **The information for each of these points is provided in the Methods, subsections on *Statistical analyses.*** |
| **Results** | | |
| Participants | 13* | (a) Report numbers of individuals at each stage of study—eg numbers potentially eligible, examined for eligibility, confirmed eligible, included in the study, completing follow-up, and analysed  **This information is provided in Figure 1.** |
|  |  | (b) Give reasons for non-participation at each stage  **This information is provided in the Methods, subsections on *Statistical analyses* and in Figure 1.** |
|  |  | (c) Consider use of a flow diagram  **This information is provided in Figure 1.** |
| Descriptive data | 14* | (a) Give characteristics of study participants (eg demographic, clinical, social) and information on exposures and potential confounders  **This information is provided in Table 1, and Supplementary Tables S2-S5** |
|  |  | (b) Indicate number of participants with missing data for each variable of interest  **This information is provided in Figure 1 and Supplementary Tables S2-S5** |
| Outcome data | 15* | Report numbers of outcome events or summary measures  **This information is provided throughout the Results and Supplementary appendices.** |
| Main results | 16 | (*a*) Give unadjusted estimates and, if applicable, confounder-adjusted estimates and their precision (eg, 95% confidence interval). Make clear which confounders were adjusted for and why they were included  **This information is provided throughout the Results and Supplementary appendices.** |
|  |  | (*b*) Report category boundaries when continuous variables were categorized  **This information is provided throughout the Results and Supplementary appendices.** |
|  |  | (*c*) If relevant, consider translating estimates of relative risk into absolute risk for a meaningful time period  **Not applicable** |
| Other analyses | 17 | Report other analyses done—eg analyses of subgroups and interactions, and sensitivity analyses  **Secondary and subgroup analyses are provided in the Results and Supplementary appendices** |
| **Discussion** | | |
| Key results | 18 | Summarise key results with reference to study objectives  **This information is provided in the Discussion, paragraph 1.** |
| Limitations | 19 | Discuss limitations of the study, taking into account sources of potential bias or imprecision. Discuss both direction and magnitude of any potential bias  **This information is provided in the Discussion, paragraph 6.** |
| Interpretation | 20 | Give a cautious overall interpretation of results considering objectives, limitations, multiplicity of analyses, results from similar studies, and other relevant evidence  **This information is provided in throughout the Discussion** |
| Generalisability | 21 | Discuss the generalisability (external validity) of the study results  **This information is provided in the Discussion, paragraph 7.** |
| **Other information** | | |
| Funding | 22 | Give the source of funding and the role of the funders for the present study and, if applicable, for the original study on which the present article is based  **This information is provided in the Funding and Acknowledgement sections.** |

*Give information separately for exposed and unexposed groups.
